# Contactless ultrasound chest vibration mapping discriminates respiratory and cardiac patients from healthy individuals

**DOI:** 10.64898/2026.05.09.26352804

**Authors:** Eric Saloux, Loïc Demore, Frédéric Wintzenrieth, Amir Hodzic, Amèle Mouadil, Mehdi Shekarnabi, Aleksey Vladimirovich Zemniskiy, Philippe Mendels-Flandre, Sam Bayat, Mathias Fink, Ros Kiri Ing, Mathieu Couade, Thomas Similowski, the AUSTRAL-AF2 investigator group

## Abstract

Contactless assessment of cardiopulmonary function remains an unmet need, with current approaches relying either on subjective clinical examination or on resource-intensive imaging. We evaluated a novel multipoint airborne ultrasound surface motion camera (SMC) designed to map thoracic vibration patterns without contact and to extract clinically relevant information through data-driven analysis.

In a prospective observational study, clinically characterised participants underwent short-duration acquisitions during natural breathing and externally induced oscillations. The resulting signals were transformed into spatially and frequency-resolved maps and analysed using machine learning models to discriminate healthy individuals from patients with respiratory or cardiac disease.

The approach proved feasible in a clinical setting and achieved excellent discrimination between healthy individuals and respiratory patients (area under the receiver operating characteristic curve (AUC) 0.90 ± 0.07), including in patients with subtle abnormalities not detected by pulmonary function testing. Discrimination between healthy individuals and cardiac patients ranged from acceptable to excellent (AUC 0.76–0.90 depending on subgroup), with the highest performance observed in aortic stenosis. Model interpretability analyses revealed spatial and spectral patterns consistent with the known physiological organisation of lung mechanics and cardiac auscultation areas, supporting a structure–function relationship between recorded signals and underlying processes.

These findings indicate that thoracic vibration transmission encodes spatially and spectrally organised information that can be captured without contact and exploited through explainable data-driven modelling. While the results require confirmation in larger populations, this approach may represent an operator-independent, low-burden extension of bedside assessment, with potential applications in early detection, triage, and monitoring of cardiopulmonary disease.

## I. Introduction

High-frequency vibrations originating within the thorax propagate to the thoracic surface, where they can be perceived by manual palpation. This constitutes one of the historical foundations of chest physical examination ^1,2^. Such vibrations can arise from high-velocity blood flow across cardiac structures or from the lungs resonating to acoustic excitation. Current medical curricula still include explicit teaching on how to detect flagrant cardiac valvular disorders through precordial thrills and on how to identify abnormalities of the lungs, pleura, and chest wall through palpation during vocalisations (tactile fremitus) ^3–6^.

Yet the palpatory assessment of thoracic vibrations has lost its operational role in contemporary clinical practice. The variability of interobserver skills is unknown but likely substantial. Interpretation is highly subjective, and operating values are unknown ^7^. The information provided is fragmentary, cannot be recorded or shared, and cannot be subjected to external validation or longitudinal comparison. In addition, direct manual contact between the examiner and the patient is mandatory.

As a result, clinicians often proceed directly to specialised investigations such as chest computed tomography, pulmonary function testing, or cardiac ultrasound, despite their costs, logistical constraints, and dependence on specialised equipment and expertise. In the case of X-ray–based techniques, imaging overuse can contribute to identified population-level serious risks, as estimated in contemporary modelling studies e.g.^8^. Default escalation to resource-intensive investigations may be particularly problematic in clinical situations where the initial cardiac or pulmonary orientation is uncertain, as it may contribute to inefficient diagnostic pathways ^9,10^. There is therefore a clear need for point-of-care, low-burden approaches that can extend and objectivise physical examination prior to external investigations. Within this framework, rapidity, limited dependence on patient cooperation, reduced operator dependence, and non-contact operation all constitute operational advantages.

Variations in the reflection of airborne acoustic waves from the thoracic surface can be used to track low-frequency motions related to the cardiac and respiratory cycles ^11,12^, based on principles derived from both sonar-based ranging and Doppler ultrasound physics. The same approach can also track high-frequency physiological phenomena. Likewise, we have previously shown in healthy individuals that a multipoint airborne ultrasound surface motion camera (SMC) can delineate cardiac velocity profiles ^13^. It can also map small-amplitude vocalisation-induced vibrations over the entire thorax and produce transmission matrices proposed to reflect structural properties of the respiratory system ^14^. This framework makes it possible to convert a traditionally qualitative clinical signal into a structured, high-dimensional spatiotemporal dataset amenable to quantitative analysis. Given the complexity and multidimensionality of these signals, data-driven modelling of their spatiotemporal structure is required to extract clinically relevant patterns. Importantly, these methods can be designed to retain physiological interpretability, allowing model-derived features to be related to underlying cardiac and respiratory mechanisms.

The present observational study was conducted to test the hypothesis that multipoint airborne ultrasound mapping using a SMC is feasible in a clinical setting, and that lung and chest wall disorders sufficiently alter thoracic vibration transmission patterns to generate discriminative, physiologically structured signals allowing discrimination between respiratory patients and healthy individuals. We also tested the hypothesis that the same technology could also discriminate cardiac patients from healthy individuals.

## II. Material and Methods

### II.1. Ethical considerations

This observational, cross-sectional, monocentric study was conducted in accordance with the principles of the Declaration of Helsinki. It was approved by the appropriate ethics committee (*Comité de Protection des Personnes* Sud-Méditerranée V, Nice, France, decision 23-0269). All participants received detailed information about the study and provided written informed consent. The study protocol had been publicly registered before the first inclusion (NCT06661200).

### II.2. Recruitment criteria

Three participant populations were studied: healthy individuals, respiratory patients, and cardiac patients. Generic inclusion criteria were age 18 years or older, clinical stability at the time of evaluation, ability to stand and to perform simple respiratory manoeuvres, and affiliation with a social security scheme. Generic exclusion criteria were a body mass index below 17 or above 35 kg·m⁻²; any condition likely to interfere with thoracic vibration acquisition (including breast implants, implanted cardiac devices, or recent thoracic surgery); known pregnancy; inability to cooperate for any reason, including major hearing or cognitive impairment; and legal guardianship. In addition, specific inclusion criteria were defined for each of the three participant populations, who underwent pulmonary function tests (PFTs) using a Jaeger MasterScreen system (CareFusion, Germany) and cardiac echocardiography using an EPIQ 7 system (Philips Healthcare, Bothell, WA, USA) equipped with an X5-1 PureWave microbeamforming xMATRIX probe.

In the healthy individuals population (Group 1), participants were recruited by word of mouth and local publicity. They had to declare themselves free of any significant medical condition and to have normal PFTs (namely forced expiratory volume in 1 s [FEV1], vital capacity [VC], total lung capacity [TLC], and corrected lung carbon monoxide transfer [TLCO] above 80% of predicted values according to Global Lung Function Initiative [GLI] equations ^15–17^) and a normal cardiac echocardiography (namely a left ventricular ejection fraction [LVEF] > 50% and no structural abnormalities ^18,19^), both performed on the study day.

In the respiratory patient population (Group 2), participants had to have been referred to the investigating centre pulmonary function testing laboratory and to exhibit either objective evidence of pulmonary disease despite normal lung function (established on clinical and/or radiological grounds) or abnormal lung function consisting in a spirometric defect with a forced expiratory volume in 1 s (FEV₁) below 80% predicted and/or a vital capacity (VC) below 80% predicted and/or a carbon monoxide transfer capacity (TLCO) below 80% predicted. Three severity subgroups were defined for sensitivity analyses: mild (FEV₁ ≥ 80%, VC ≥ 80%, and TLCO ≥ 80% but established pulmonary disease; Group 2-1), moderate (FEV₁ or VC or TLCO 50–80%; Group 2-2), and severe (FEV₁ or VC or TLCO < 50%; Group 2-3).

In the cardiac patient population (Group 3), participants had to have been referred to the investigating centre cardiac ultrasound laboratory and to have received a diagnosis of: (1) moderate-to-severe aortic stenosis (aortic valve area ≤ 1.5 cm² by the continuity equation ^19^; Group 3-1); (2) moderate-to-severe aortic regurgitation (effective regurgitant orifice area [EROA] ≥ 20 mm², regurgitant volume [RVol] ≥ 45 mL·beat⁻¹, regurgitant fraction [RF] > 50% ^19,20^; Group 3-2); (3) moderate-to-severe mitral regurgitation (EROA ≥ 30 mm², RVol ≥ 45 mL·beat⁻¹, RF > 50% ^19,20^; Group 3-3); (4) hypertrophic or infiltrative cardiomyopathy (Group 3-4); or (5) dilated cardiomyopathy of ischaemic or other origin (Group 3-5). In Groups 3-4 and 3-5, left ventricular remodelling was assessed using linear measurements and derived indices, in accordance with American Society of Echocardiography recommendations ^18^. Participants presenting cardiac abnormalities compatible with more than one subgroup were not included.

### II.3. Measurement technology and raw data acquisition

The technology used in this study has been described in detail elsewhere ^14^. In brief, the SMC is a non-contact airborne ultrasound system designed for wide-field mapping of thoracic and abdominal surface vibrations with high temporal resolution. An antenna placed 50–70 cm in front of or behind the examined person and covering an area of approximately 30 × 40 cm comprises a wide-aperture ultrasonic transmitter array with 960 emitting elements, which generates broadband linear chirps spanning 30–60 kHz. Each chirp corresponds to a transmission frame whose duration equals the inverse of the pulse repetition frequency (PRF), which can reach up to 1 kHz. Airborne echoes returning from the thoracic surface are recorded synchronously by a 16 × 16 microphone reception array (256 channels). It allows chest movements along the ultrasound beam axis to be measured at multiple locations. For each transmission, it yields a time series of acoustic channel raw data, in which the samples encode echo amplitude and time-of-flight (TOF) variations related to surface motion. In parallel, an RGB-depth camera (Intel RealSense D415) captures the three-dimensional external geometry of the thorax, providing a dense surface grid used as the spatial support for motion reconstruction.

### II.4. Data acquisition protocol

#### II.4.1. Cardiac acquisitions

Cardiac acquisitions were performed with the participants standing upright and facing the antenna panel. A three-lead ECG was recorded simultaneously to provide precise R-peak timing. Each participant underwent four SMC recordings, two ∼10 s free breathing and two ∼7 s during a brief breath-hold.

#### II.4.2. Pulmonary acquisitions

Pulmonary acquisitions were performed with participants standing upright, with their back facing the antenna panel. Participants breathed quietly for a minute through a mouthpiece connected to an oscillometry device (RESMON Pro®, Restech Srl, Milan, Italy), with a nose clip in place and the cheeks supported. Ten-second SMC recordings were performed synchronously with forced oscillatory stimulation superimposed on natural breathing cycles.

### II.5 Signal processing and data analysis

#### II.5.1. Pre-processing and motion reconstruction

From the raw acquisitions described above, the internal signal-processing pipeline, previously described in detail ^14^, can be summarised as follows. The thoracic surface is first reconstructed using the RGB-depth camera, providing a three-dimensional surface map with millimetric accuracy. This map provides the spatial coordinates of each point of the thorax, which are then used to compute the receive delays required to focus the array at each location according to a predefined delay law. Using these precomputed delays, the reception matrix is numerically focused through a software beamforming procedure based on time-domain delay-and-sum. For each point of the reconstructed surface, the signals received by all array elements are delayed accordingly and coherently summed to reconstruct a focused temporal signal representative of the local acoustic response. This operation is performed for each acquisition frame, yielding a sequence of focused surface maps sampled at the pulse repetition frequency (PRF). Inter-frame motion is subsequently estimated by computing phase shifts between consecutive frames using a cross-correlation approach adapted to airborne ultrasound. These phase shifts are converted into surface velocity estimates, providing a spatiotemporal representation of thoracic vibration dynamics.The resulting SMC pipeline achieves temporal imaging rates of up to 1 kHz, a spatial resolution of approximately 3 cm, and velocity noise on the order of 100 µm·s⁻¹ ^14^.

In the present study, cardiac acquisitions were performed with an imaging rate set to about 244Hz (master clock at 1 MHz, clock divider set to 4096), whereas pulmonary acquisitions were performed with an imaging rate set to about 122 Hz (master clock at 1 MHz, clock divider set to 8192) with a maximum excitation frequency of 37 Hz. Acquisition durations ranged from 7 s for breath-held cardiac acquisitions to 10 s for free breathing and respiratory oscillometry acquisitions. From this point onward, specific processing pathways are followed to analyse surface manifestations of cardiac activity and the transmission of lung-related vibrations (**Figure 1A**).

**Figure 1.**
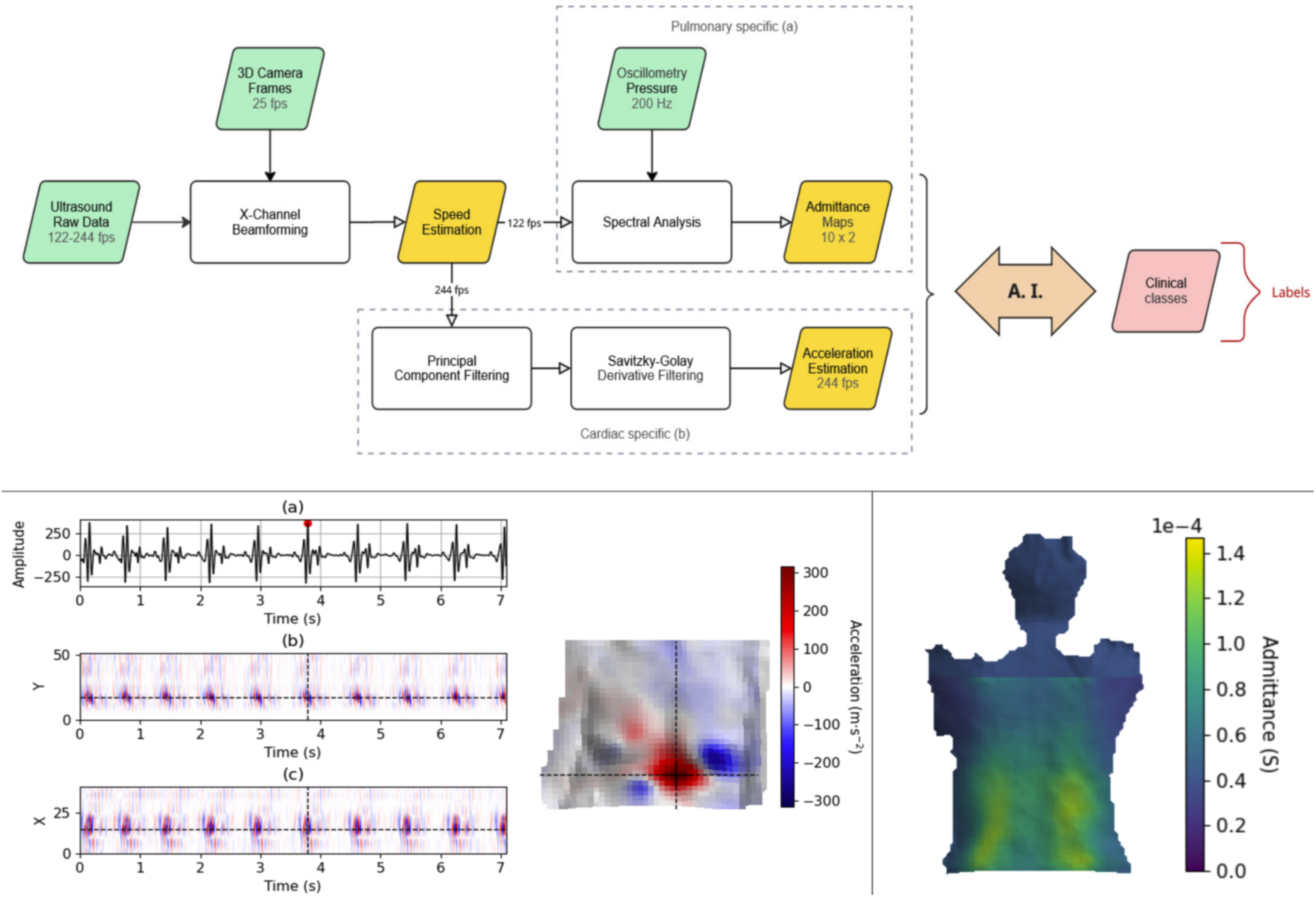
Data processing pipelines. **A. Overview of the data processing pipeline** converting ultrasound raw data into SMC-C and SMC-L outputs for AI-based analysis. **B. Visualisation of the cardiac SMC modality.** *(a) Temporal seismocardiographic (SCG) waveform extracted at a fiducial point on the chest surface (black marker), selected as the location of maximal intensity across spatial and temporal dimensions of the SMC-C data. (b) Vertical slice and (c) longitudinal slice of the SCG volume, shown as temporal maps. (d) Spatial SCG slice projected onto the three-dimensional surface reconstruction of a patient’s thorax, illustrating the distribution of cardiac-related surface vibrations.* **C. Visualisation of the pulmonary SMC modality.** *Admittance map at a selected excitation frequency projected onto the three-dimensional surface of a patient’s back*.

Regarding cardiac activity (SMC-C), surface velocity signals are converted into seismocardiograms by removing the first singular vector obtained via singular value decomposition (SVD), which corresponds to global displacement and breathing-related motion. The resulting signal is then processed with an infinite impulse response (IIR) bandpass filter and subsequently converted to surface acceleration using a Savitzky–Golay finite impulse response (FIR) filter ^21^. The final outputs of this pipeline consist of three-dimensional vibration volumes (N × 41 × 51), where N denotes the total number of frames per acquisition (N = 1732 for acquisitions performed during breath-holding and N = 2500 for acquisitions performed during free breathing). **Figure 1B** summarises this process visually. These representations are used directly as inputs to the “cardiac” deep-learning models (**Figure 1B**).

Regarding surface transmission of pulmonary vibrations (SMC-L), a Fast Fourier Transform (FFT) is applied to each spatial channel of the oscillometry recordings to generate frequency-resolved admittance maps. For each spatial location, complex admittance Y(f) is computed as the ratio between measured surface velocity and applied oscillatory pressure. Admittance quantifies how efficiently oscillatory pressure is converted into thoracic surface motion (mechanical transfer function) and is expressed as Y(f) = G(f) + jB(f), where the real component G(f) (conductance) reflects dissipative mechanical behaviour, and the imaginary component B(f) (susceptance) reflects the elastic and inertial energy storage properties of the respiratory system. This process yields 20 aligned two-dimensional maps corresponding to the real and imaginary components at 10 excitation frequencies contained in the pseudorandom signal delivered by the oscillometry system described above (5, 7, 11, 13, 17, 19, 23, 29, 31, and 37 Hz sequence) (**Figure 1C**). These maps constitute the input to the “pulmonary” deep-learning models.

### II.5.2. Deep-learning models

#### II.5.2.1. Input representations

On the basis of the internal SMC reconstruction pipeline described above, no additional signal-processing steps were performed prior to model input. Cardiac recordings were thus represented as calibrated three-dimensional spatiotemporal volumes corresponding to approximately 7 s (breath-holding) to 10 s (free breathing) of recording. Pulmonary recordings were represented by a set of 20 spatial admittance maps derived from the forced-oscillation stimulus. After quality checks, only one recording (instead of two) was available for analysis in 7.3%, 19.8%, and 19.4% of participants retained for algorithm training in the apnoea, free-breathing, and oscillometry conditions, respectively.

#### II.5.2.2. Cardiac model

The cardiac classifier (**Figure S1A**) consisted of a three-dimensional convolutional neural network receiving input tensors of size N × 41 × 51. The architecture included two 3D convolutional blocks with batch normalisation and 3D max pooling, followed by a time-distributed flattening operation, two bidirectional long short-term memory (LSTM) layers, and two dense layers with dropout regularisation. A final SoftMax layer provided the probability of cardiac abnormality versus healthy status. Training was performed with a learning rate of 8 × 10⁻⁶, a batch size of 8, a dropout ratio of 0.5, and 200 epochs on NVIDIA L40S or H100 GPUs, using categorical cross-entropy.

#### II.5.2.3. Pulmonary model

The pulmonary classifier (**Figure S1B**) was similar to the cardiac model, except that the LSTM layers were replaced by a one-dimensional flattening operation across all dimensions. Training used the same hyperparameters as described above.

#### II.5.2.4. Validation strategy

Both models were trained and evaluated using a five-fold cross-validation scheme applied in a strictly subject-wise manner, such that all recordings from a given participant were assigned exclusively to either the training or the validation set, thereby preventing data leakage. For each fold, dataset partitioning was stratified to preserve group proportions across training and validation splits. The entire procedure was repeated five times with different random partitions to enhance statistical reliability. Performance was assessed separately for the cardiac and pulmonary tasks.

#### II.5.2.5. Experiments

Fourteen experiments were conducted to evaluate the models’ ability to discriminate Group 1 participants from Group 2 and Group 3 participants. Ten training experiments addressed Group 3 subpopulations, including all heart disease (Group 3), valvular heart disease (Group 3-1, 3-2 and 3-3), muscular heart disease (Group 3-4 and 3-5), aortic stenosis (Group 3-1), mitral regurgitation (Group 3-3) each analysed separately according to acquisition type (breath-holding and free breathing) Four additional training experiments addressed Group 2 participants, including all lung disease (Group 2), mild lung disease (Group 2-1), moderate lung disease (Group 2-2) and severe lung disease (Group 2-3).

### II.5.3. Explainability

Model interpretability was evaluated using Gradient-weighted Class Activation Mapping (Grad-CAM). Gradients of the predicted class score were computed with respect to the activations of the final convolutional layer, yielding a three-dimensional tensor of shape (D, W, H), where W and H denote spatial dimensions, and D corresponds to the temporal dimension for cardiac data or the frequency dimension for pulmonary data. This approach was used to provide a global explanation of the features contributing to the models’ decisions. Three complementary analyses were performed to provide temporal, frequential, and spatial interpretations of the models’ decisions. For temporal analysis, synchronised ECG data served as the reference. The three-dimensional Grad-CAM tensor was first reduced to a one-dimensional temporal activation signal by maximum projection along the spatial dimensions. To assess alignment between model attention and cardiac phase, peaks were detected in both the gradient-derived activation signal and the ECG signal (specifically R-peaks). Temporal consistency was then quantified by calculating the number of gradient peaks falling within a tolerance window of width L centred on each ECG R-peak. For frequential analysis (pulmonary model), the spatial dimensions were reduced by maximum projection to obtain a one-dimensional activation signal along the excitation frequency dimension.Frequential consistency was then assessed through comparison between control and patient groups. For spatial analysis, activation maps were mean-projected onto the thoracic surface reconstructed from the depth camera and averaged across acquisitions, allowing spatial interpretation of the regions most influential in the classification decision. Attention patterns were inspected to assess their consistency with physiological expectations, such as predominant upper–anterior activation for cardiac recordings and posterior–basal activation for pulmonary recordings.

### II.6 Model performance metrics

Model outputs were expressed as posterior class probabilities and evaluated using receiver operating characteristic (ROC) analyses. For each validation fold, ROC curves were constructed by varying the decision threshold applied to the model-derived probabilities and plotting sensitivity against 1–specificity. The area under the ROC curve (AUC) was used as the primary summary measure of discriminative performance. Additional metrics, including sensitivity, specificity, accuracy and F1 score, were computed on the validation data using a fixed decision threshold of 0.5 applied to the predicted class probabilities. For descriptive purposes, optimal sensitivity and specificity values were also derived from the ROC curves using the Youden index (positive and negative predictive values were not reported, as these metrics depend on disease prevalence and are only interpretable in clinically representative populations with a defined prevalence structure). When multiple recordings were available for a given participant, recording-level outputs were aggregated to yield one value per participant prior to final performance estimation. Intergroup comparisons of ROC curves were performed using the Kruskal–Wallis test to assess whether one group stochastically outperformed the others; in the event of a significant result, pairwise comparisons were conducted using Dunn’s test with Bonferroni correction. Differences in secondary performance metrics (sensitivity, specificity, accuracy, and F1 score) were assessed using the Kruskal–Wallis test applied to fold-level estimates.

## III. Results

### III.1. Study populations and data availability

A total of 227 participants were included in the study over the protocol pre-specified duration of 12 months (May 2024–April 2025). Ten participants were secondarily excluded because of violations of the inclusion criteria. Among the remaining 217 participants retained for population analysis, 33 belonged to Group 1 (**Table 1**), 87 to Group 2 (**Table 2**), and 97 to Group 3 (**Table 3**). Among these, four participants in Group 2 were unable to undergo preliminary oscillometry testing, resulting in SMC recordings being available for 213 participants across all acquisition modalities. Some acquisitions were later excluded from algorithm modelling following a series of 20 technical and quality-control checks identifying issues such as low signal-to-noise ratio, dead channels, electrocardiogram Q-peak detection failure, oscillometry dysfunction, etc. (**Table 4**). In addition, one case of mild respiratory condition was excluded because of the absence of radiological abnormalities. The final modelling therefore pertained to 198 individuals: 33 in Group 1, 71 in Group 2, and 94 in Group 3. Subgroup 3-4 (hypertrophic or infiltrative cardiomyopathy) proved difficult to populate owing to the high prevalence of cardiac pacemakers and defibrillators and was therefore merged with subgroup 3-5 (dilated cardiomyopathy of ischaemic or other origin) for analysis. The characteristics of the patients according to subgroups are presented in the electronic supplement (**Table S1** and **Table S2**). In Groups 2 and 3, patients were included on a consecutive basis, modulated by routine workload and personnel availability.

**Table 1.** Characteristics of the healthy participants included in the study (n = 33).

|  |  |
| --- | --- |
| Sex (W/M) | **16/17** |
| Age (years) | **52.0** [26.0 - 63.0] (18.0-80.0) |
| Height (cm) | **170.0** [167.0 - 173.0] (151.0 - 187.0) |
| Weight (kg) | **69.0** [63.0 - 76.0] (52.0 - 100.0) |
| Body mass index (kg.m<sup>-2</sup>) | **24.1** [21.6 - 26.7](18.1 - 34.6 ) |
| FEV1 (L) | **3.7** [2.8 - 4.0] (1.9 - 5.4) |
| FEV1 (% pred) | **110.0** [99.0 - 118.0] (84.0 - 147.0) |
| VC (L) | **4.5** [3.9 - 5.3] (2.6 - 7.0) |
| VC (% pred) | **124.0** [105.0 - 131.0 ] (82.0 - 158.0) |
| LVEF (%) | **64.0** [61.0 - 68.0] (55.0 - 75.0) |

Data are presented as median [first quartile - third quartile](minimum-maximum). FEV1: forced expiratory volume in 1s; VC: vital capacity; % pred: percentage of predicted values according to the Global Lung Function Initiative [GLI] equations; LVEF : left ventricular ejection fraction.

**Table 2.**
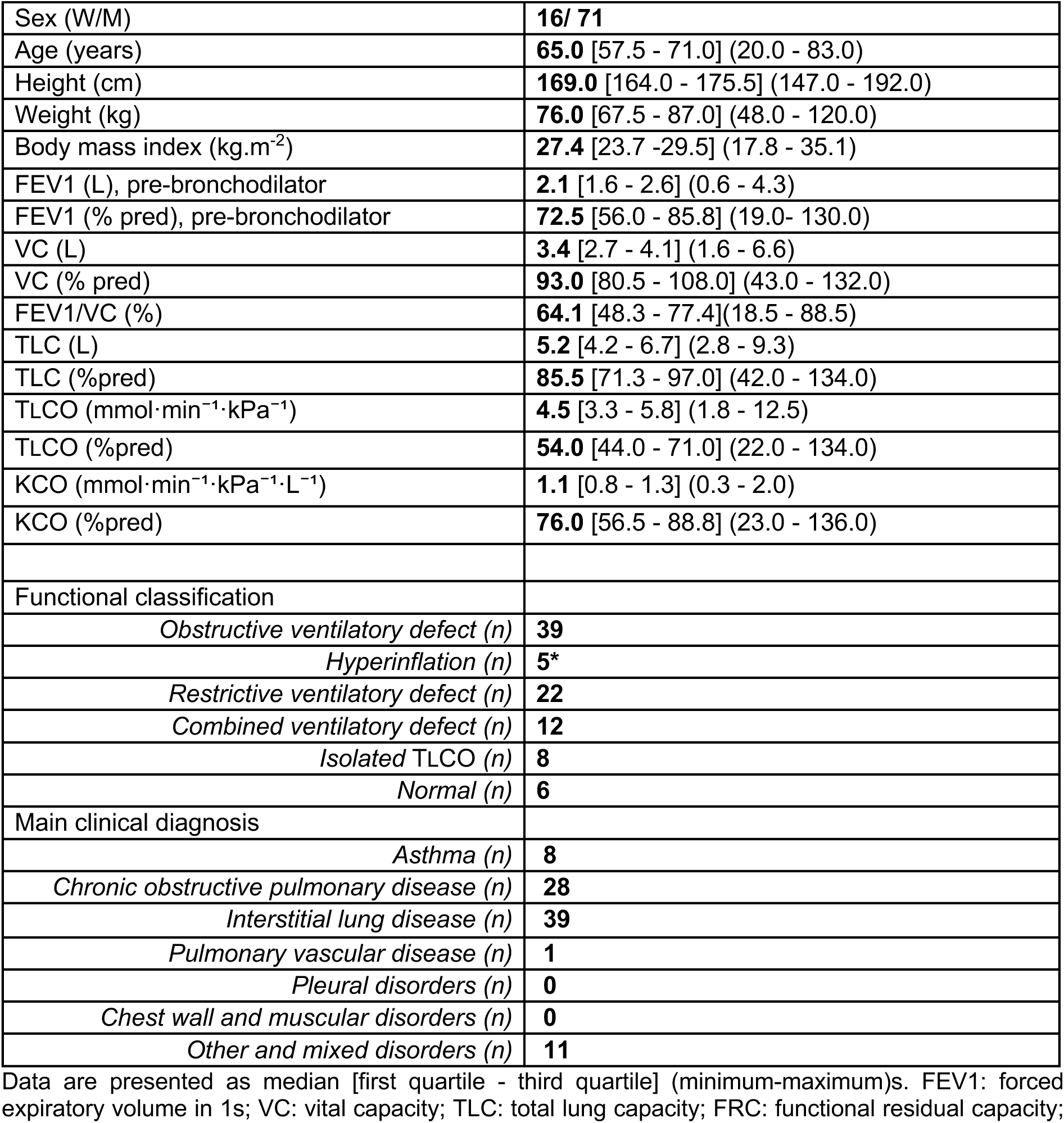
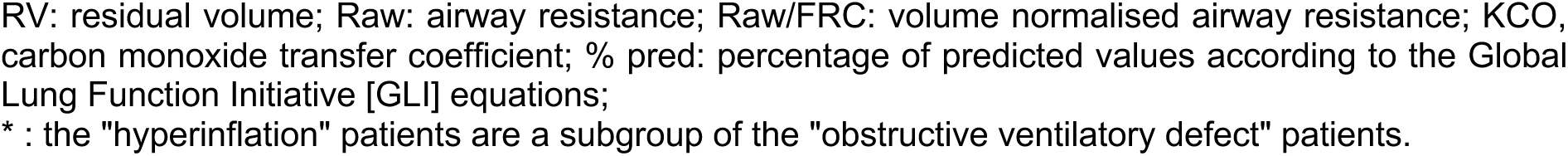
Characteristics of the respiratory patients included in the study (n = 87).

|  |  |
| --- | --- |
| Sex (W/M) | **16/ 71** |
| Age (years) | **65.0** [57.5 - 71.0] (20.0 - 83.0) |
| Height (cm) | **169.0** [164.0 - 175.5] (147.0 - 192.0) |
| Weight (kg) | **76.0** [67.5 - 87.0] (48.0 - 120.0) |
| Body mass index (kg.m<sup>-2</sup>) | **27.4** [23.7 -29.5] (17.8 - 35.1) |
| FEV1 (L), pre-bronchodilator | **2.1** [1.6 - 2.6] (0.6 - 4.3) |
| FEV1 (% pred), pre-bronchodilator | **72.5** [56.0 - 85.8] (19.0- 130.0) |
| VC (L) | **3.4** [2.7 - 4.1] (1.6 - 6.6) |
| VC (% pred) | **93.0** [80.5 - 108.0] (43.0 - 132.0) |
| FEV1/VC (%) | **64.1** [48.3 - 77.4](18.5 - 88.5) |
| TLC (L) | **5.2** [4.2 - 6.7] (2.8 - 9.3) |
| TLC (%pred) | **85.5** [71.3 - 97.0] (42.0 - 134.0) |
| TLCO (mmol·min<sup>-1</sup>·kPa<sup>-1</sup>) | **4.5** [3.3 - 5.8] (1.8 - 12.5) |
| TLCO (%pred) | **54.0** [44.0 - 71.0] (22.0 - 134.0) |
| KCO (mmol·min<sup>-1</sup>·kPa<sup>-1</sup>·L<sup>-1</sup>) | **1.1** [0.8 - 1.3] (0.3 - 2.0) |
| KCO (%pred) | **76.0** [56.5 - 88.8] (23.0 - 136.0) |
| Functional classification |  |
| Obstructive ventilatory defect (n) | **39** |
| Hyperinflation (n) | **5\*** |
| Restrictive ventilatory defect (n) | **22** |
| Combined ventilatory defect (n) | **12** |
| Isolated TLCO (n) | **8** |
| Normal (n) | **6** |
| Main clinical diagnosis |  |
| Asthma (n) | **8** |
| Chronic obstructive pulmonary disease (n) | **28** |
| Interstitial lung disease (n) | **39** |
| Pulmonary vascular disease (n) | **1** |
| Pleural disorders (n) | **0** |
| Chest wall and muscular disorders (n) | **0** |
| Other and mixed disorders (n) | **11** |

Data are presented as median [first quartile - third quartile] (minimum-maximum)s. FEV1: forced expiratory volume in 1s; VC: vital capacity; TLC: total lung capacity; FRC: functional residual capacity;
RV: residual volume; Raw: airway resistance; Raw/FRC: volume normalised airway resistance; KCO, carbon monoxide transfer coefficient; % pred: percentage of predicted values according to the Global Lung Function Initiative [GLI] equations;
\* : the "hyperinflation" patients are a subgroup of the "obstructive ventilatory defect" patients.

**Table 3.**
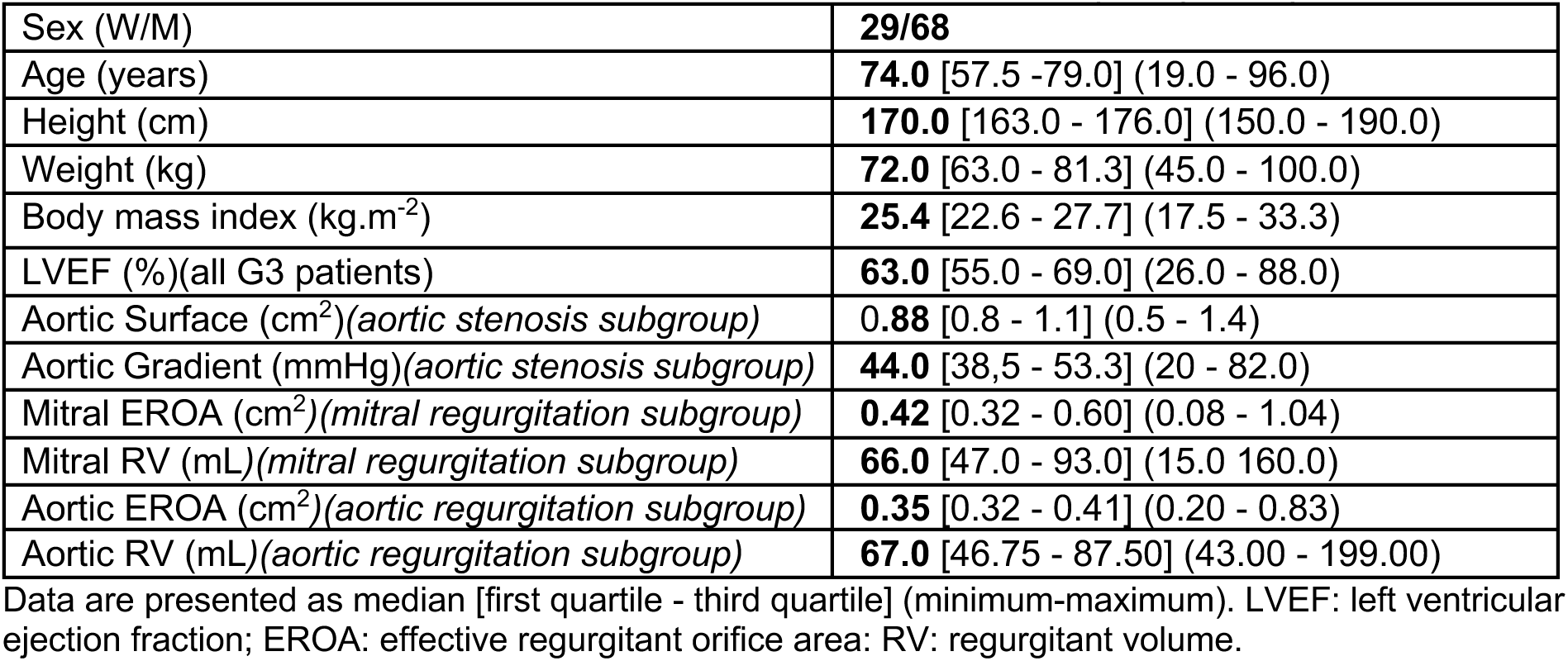
Characteristics of the cardiac patients retained for analysis (n = 97).

**Table 4.**
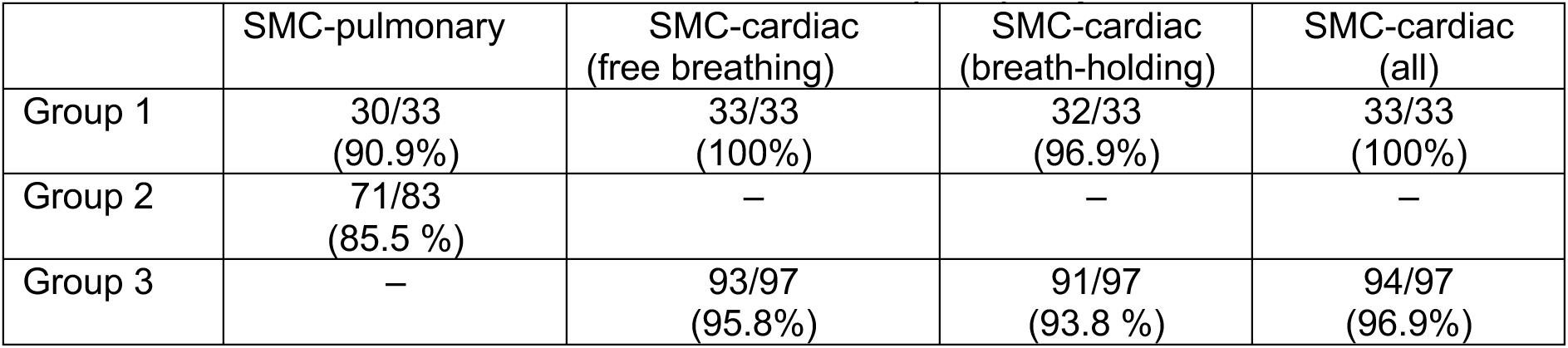
Rates of successful surface motion camera (SMC) acquisitions across modalities.

|  | SMC-pulmonary | SMC-cardiac<br>(free breathing) | SMC-cardiac<br>(breath-holding) | SMC-cardiac<br>(all) |
| --- | --- | --- | --- | --- |
| Group 1 | 30/33<br>(90.9%) | 33/33<br>(100%) | 32/33<br>(96.9%) | 33/33<br>(100%) |
| Group 2 | 71/83<br>(85.5 %) | — | — | — |
| Group 3 | — | 93/97<br>(95.8%) | 91/97<br>(93.8 %) | 94/97<br>(96.9%) |

### III.2. Feasibility and acceptability

Respiratory- and cardiac-related surface vibration maps were consistently acquired across the different populations studied. Examples of acquisitions are provided in the electronic supplementary materials (**Videos S1–S5**). Performing a respiratory acquisition required approximately one minute in addition to the routine pulmonary function testing procedure, compared with 20-30 seconds added to the routine cardiac ultrasound procedure for cardiac acquisitions. The rate of successful acquisition ranged from 85.0% (in Group 2) to 100 % (for cardiac acquisitions in Group 1 (**Table 4**). Acquisition failures were due to inability to perform oscillometry (independent of SMC), oscillometry–SMC synchronisation failure, prototype hardware failure, or absence of oscillometry recording.

Overall satisfaction with the study was reported by 99.5% of participants. Image acquisition with the SMC device was considered comfortable in 98.1% of questionnaires, and removal of upper-body clothing was not regarded as a source of discomfort in 95.2%. Breath-holding was well tolerated by 98.6% of participants, including repeated breath-holds (97.6%). Comfort during the acquisition procedure was reported to be similar for anterior and posterior recordings by 89.5% of the study population. During respiratory acquisitions, breathing through the RESMON Pro device was considered comfortable by 85.6% of participants. The fact that the examination was performed by a machine rather than by a human operator did not affect participants in 91.4% of cases. Most participants (96.7%) considered that the acquisition time was acceptable in a doctor’s office, and 99.5% indicated that they would recommend the experience to friends or relatives.

### III.3. Discrimination between respiratory patients and healthy individuals

#### III.3.1. Respiratory SMC-based model performance

The performance of the SMC approach in discriminating respiratory patients from healthy individuals is shown in **Figure 2**. Significant intergroup differences were observed for AUC, specificity, and F1 score (**Table S3**). Post hoc pairwise comparisons revealed significant differences in AUC between Group 1 and Group 2 (p = 0.014) and between Group 1 and Group 2-2 (p = 0.012) (**Table S3**).

**Figure 2.**
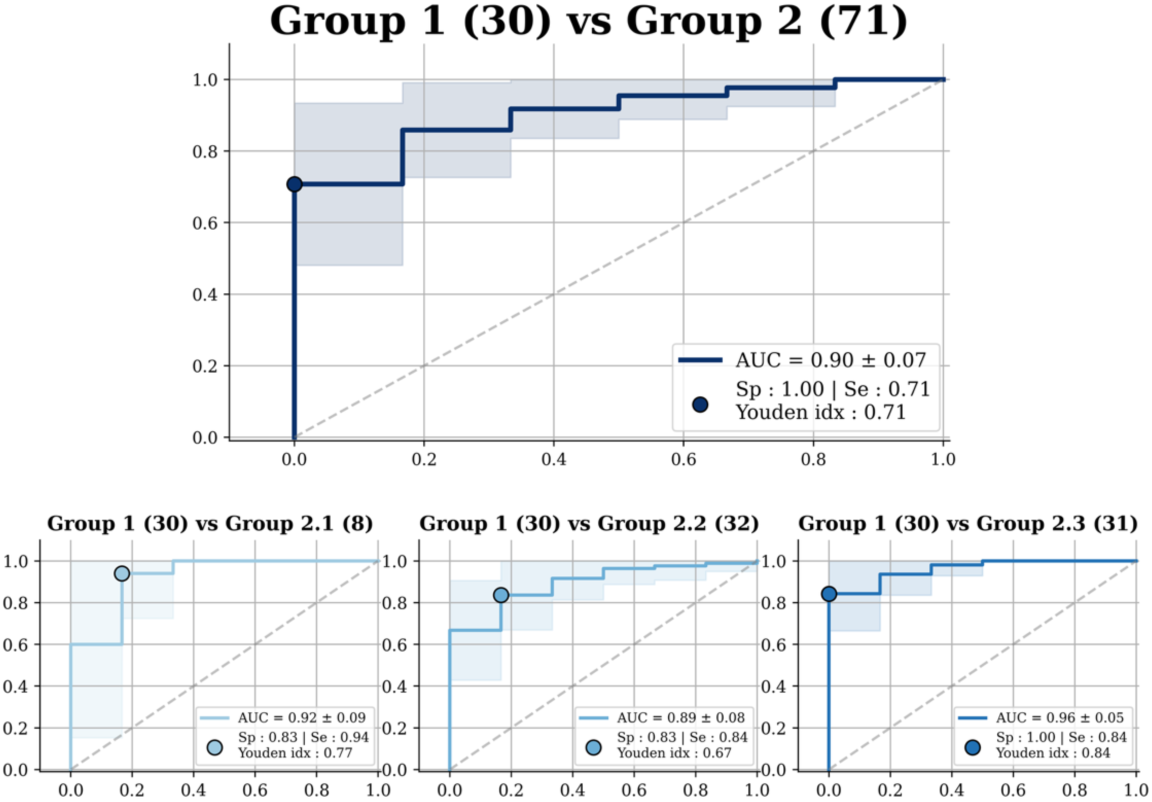
Performance of the SMC to separate healthy participants from respiratory patients. The top panel displays the global training results. The bottom panels show a sensitivity analysis by severity level: Group 1 vs. Group 2-1 (left), Group 1 vs. Group 2-2 (middle), and Group 1 vs. Group 2-3 (right).

#### III.3.2. Respiratory SMC-based model explainability

Group-averaged admittance profiles demonstrated clear frequency-dependent differences between Group 1 and Group 2-3 (**Figure 3**). In Group 1, conductance exhibited a structured frequency profile characterised by a relative peak in the low-to-mid frequency range followed by progressive attenuation at higher frequencies. In contrast, Group 2-3 patients displayed a flatter profile with reduced low and mid frequency prominence and altered high-frequency behaviour (**Figure 3A**). Similar separation was observed for susceptance, with Group 2-3 patients showing a modified frequency slope and reduced magnitude variation across excitation frequencies. On the susceptance curves, the sign reversal (zero crossing) occurred around 7 Hz in Group 1 and around 17 Hz in Group 2, indicating a shift toward higher resonance frequencies of the respiratory system in the respiratory patients (**Figure 3B**). Group-averaged Grad-CAM analysis showed that model attention along the excitation frequency dimension was preferentially concentrated in frequency bands corresponding to the largest inter-group differences in admittance as well as in regions showing the most marked signal alterations. Frequencies that most clearly separated Group 1 and Group 2-3 exhibited consistently higher gradient magnitudes, suggesting that the classifier relied on stable, disease-related redistribution of frequency-dependent mechanical transmission. Spatially resolved analyses further indicated that these frequency-dependent alterations were not uniformly distributed across the thoracic surface (**Figure 3C**). Spatial representations of the conductance and susceptance maps revealed structured spatial–frequency patterns, with Group 2-3 patients exhibiting regionally attenuated low-frequency dominance in terms of conductance and altered high-frequency attenuation in terms of susceptance. Regions displaying the greatest inter-group differences overlapped with areas identified by Grad-CAM as contributing most strongly to classification. Gradient distributions were predominantly concentrated in the lower central portion of the posterior thoracic surface and showed strong overlap with regions of highest signal coverage, indicating that model attention was aligned with physiologically and technically relevant areas **(Figure 4A).** These findings support the physiological coherence of the pulmonary model at the group level.

**Figure 3.**
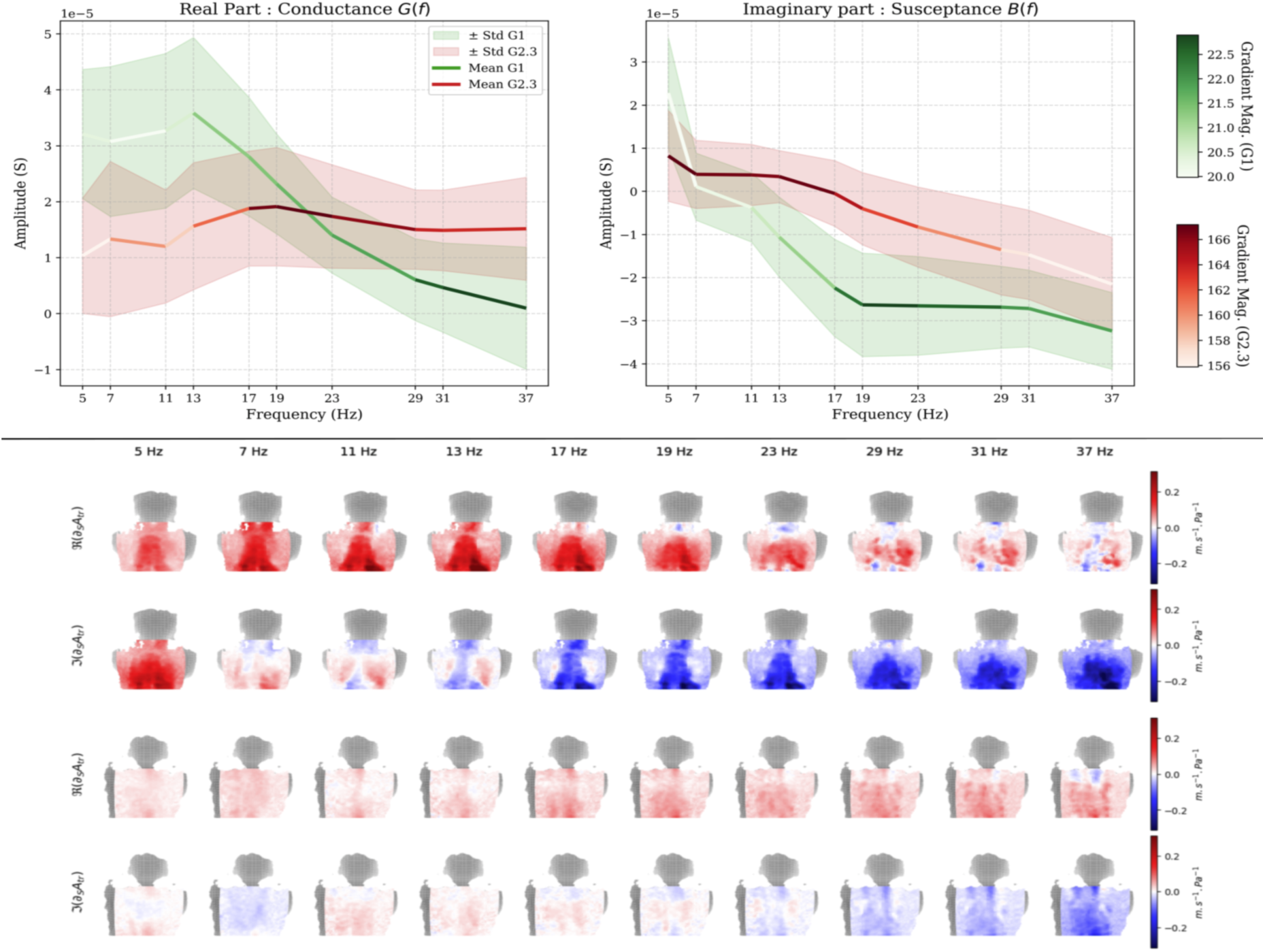
Respiratory discrimination: frequency-domain admittance and model attention. **A.** Group-averaged conductance as a function of excitation frequency in healthy participants (Group 1) and patients with severe respiratory impairment (Group 2-3). **B.** Group-averaged susceptance across the same frequency range. In A and B, curves are coloured according to the magnitude of the Grad-CAM–derived gradients along the excitation frequency dimension, reflecting the relative contribution of each frequency band to model classification. Frequencies showing the largest inter-group differences correspond to higher gradient magnitudes. **C.** Spatial representations of conductance (*R*(∂ₛA_tr_)) and susceptance *I* (I(∂ₛA_tr_)) across excitation frequencies projected onto the three-dimensional posterior thoracic surface. Columns correspond to the excitation frequencies (5–37 Hz). The two upper rows show Group 1 (healthy individuals), and the two lower rows show Group 2-3 (severe respiratory patients). Colour intensity reflects admittance magnitude. Compared with Group 1, Group 2-3 patients exhibit structured regional modifications of frequency-dependent admittance. Regions displaying the largest inter-group differences overlap with areas identified by Grad-CAM as most influential for model classification.

**Figure 4.**
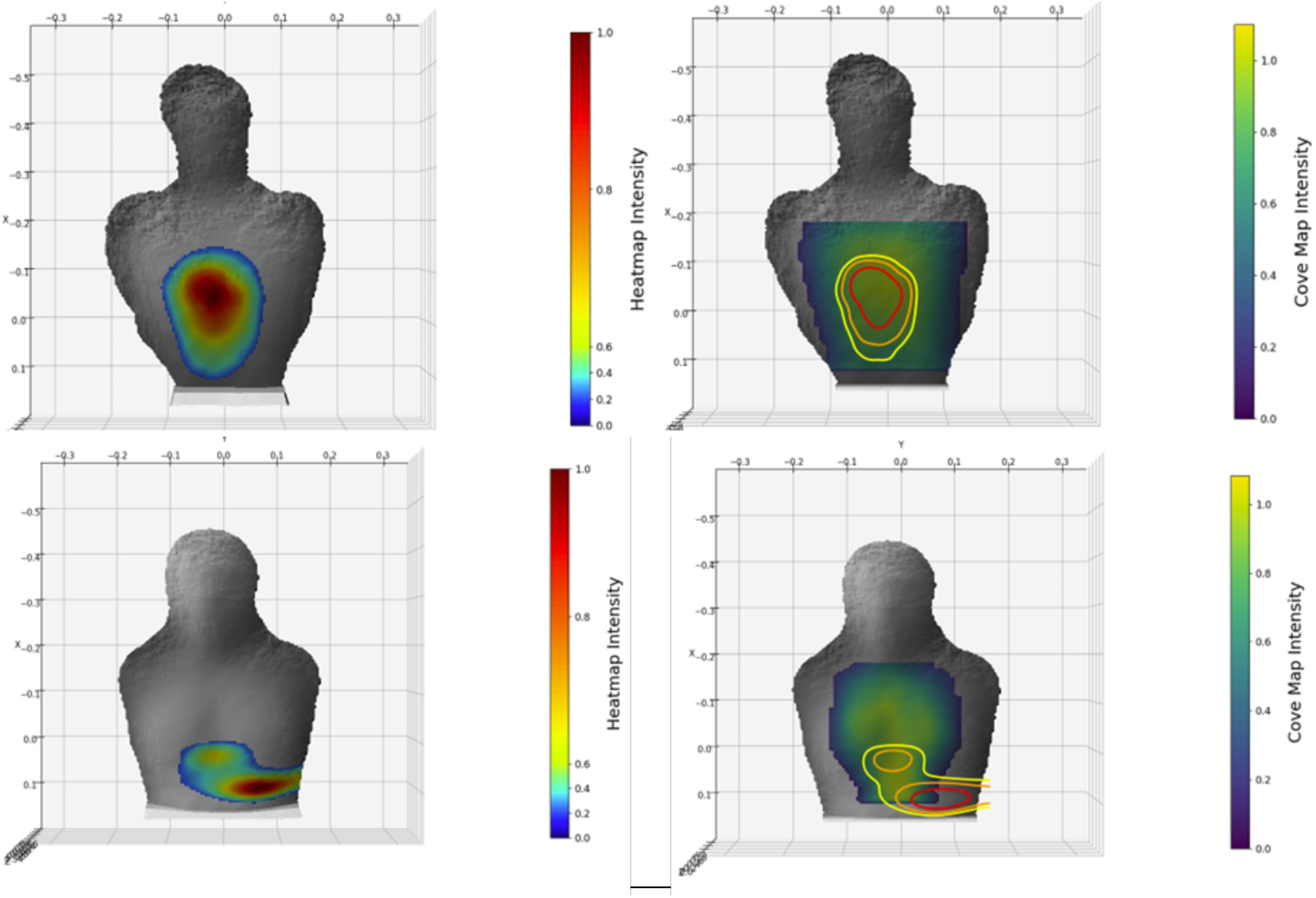
Representative thoracic surface profiles and model gradient distributions in respiratory and cardiac conditions. Regions of highest gradient intensity are predominantly located in the lower left and central thoracic regions. **Top row.** Representative three-dimensional surface profiles and gradient distributions in the respiratory condition. Left: mean-projected gradient heatmaps. Right: gradient isocontours (70–90%) overlaid on mean signal coverage maps. **Bottom row.** Same representation in the cardiac condition. Left: mean-projected gradient heatmaps. Right: gradient isocontours (70–90%) overlaid on mean signal coverage maps.

### III.4. Discrimination between cardiac patients and healthy individuals

#### III.4.1. Cardiac SMC-based model performance

The performance of the SMC approach in discriminating cardiac patients from healthy individuals is illustrated in **Figure 5 (**cardiac acquisitions performed during breath-holding). Significant intergroup differences in AUC were observed in both acquisition conditions (apnoea: H = 23.8, p = 8.7 × 10⁻⁵; free breathing: H = 28.3, p = 1.1 × 10⁻⁵) (**Table S4** and **Table S5**). Post-hoc pairwise comparisons revealed significant differences in AUC between Group 1 and Group 3.1 and between Group 1 and Group 3 (p = 2.3 × 10⁻⁴), Group 1 and Group 3-[1–2–3] (p = 1.9 × 10⁻³), Group 1 vs. Group 3-[4–5] (p = 6.3 × 10⁻⁵), and Group 1 and Group 3-3 (p = 3.4 × 10⁻⁴) in the free-breathing condition (**Table S4)**. In the apnoea condition, significant differences in AUC were also observed between Group 1 and Group 3.1 and between Group 1 and Group 3 (p = 6.3 × 10⁻³), Group 1 and Group 3-[4–5] (p = 1.9 × 10⁻⁴), and Group 1 vs. Group 3-3 (p = 8.2 × 10⁻⁴) (Table **Table S5)**.

**Figure 5.**
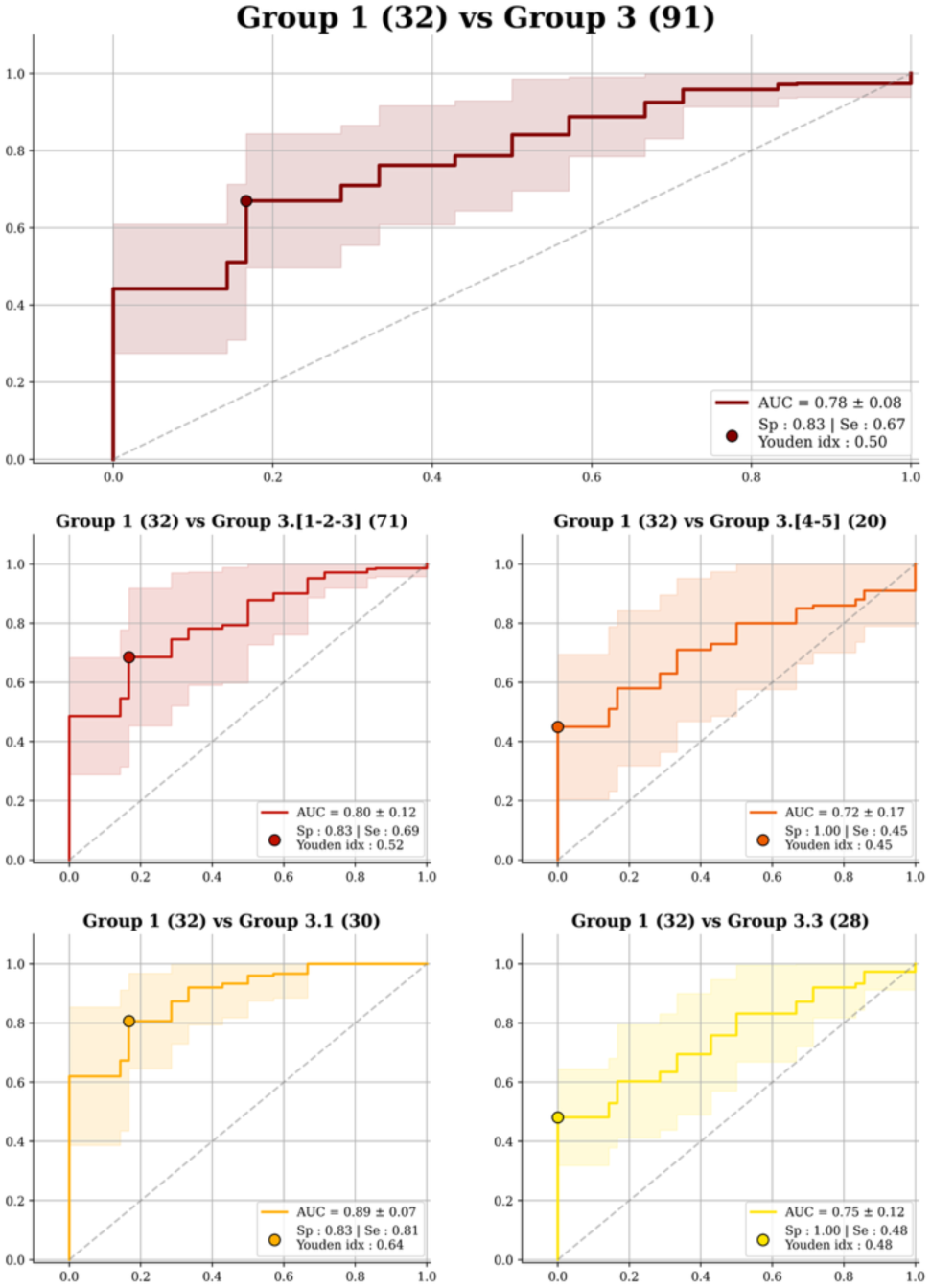
ROC curves describing the performance of the SMC-based model under apnoea conditions in discriminating cardiac patients from healthy participants. The top panel displays the global training results. The middle panels show sensitivity analyses by disease category: Group 1 vs. Group 3-[1–3] (valvular heart disease) (left) and Group 1 vs. Group 3-[4–5] (cardiomyopathies) (right). The bottom panels show sensitivity analyses for the most populated and clinically frequent subgroups: Group 1 vs. Group 3-1 (left) and Group 1 vs. Group 3-3 (right).

#### III.4.2. Cardiac SMC-based model explainability

Spatial analysis showed structured gradient patterns, with maximal intensities predominantly located in the lower left and central regions of the average three-dimensional thoracic surface (**Figure 4B**). These regions correspond to areas of highest cardiac-related vibration activity in the SMC signal. The distribution of gradient intensities also showed partial overlap with the mean antenna coverage area, indicating that model-relevant features were preferentially located within regions effectively sampled during acquisition.

Temporal analysis showed close alignment between ECG R-peaks and gradient maxima within a tolerance window of 0.3 s (**Figure 6A**). In representative recordings, detected ECG R-peaks closely matched temporal gradient peaks within this window, in some cases yielding a synchronisation score of 1.0. A slight offset between ECG peaks and gradient maxima was consistently observed. Temporal consistency analysis showed that synchronisation scores between gradient maxima and ECG signals remained high across validation participants. The score calculation was applied to all validation patients across the repeated five-fold cross-validation procedure in order to reflect the overall temporal trend. Score distributions were right-skewed, with high central and upper quantiles. For example, in the classification of Group 1 versus valvular heart disease (Group 3-[1–2–3]) during breath-hold acquisitions, the distribution showed a median synchronisation score of 0.89 (89%), a 70th percentile of 0.95 (95%), and a 90th percentile of 1.00 (100%) (**Figure 6B**). Across classification tasks, more than 50% of model inferences yielded synchronisation scores between 0.75 and 0.90. Comparable distributions were observed across other classification tasks and acquisition conditions (**Table S6**).

**Figure 6.**
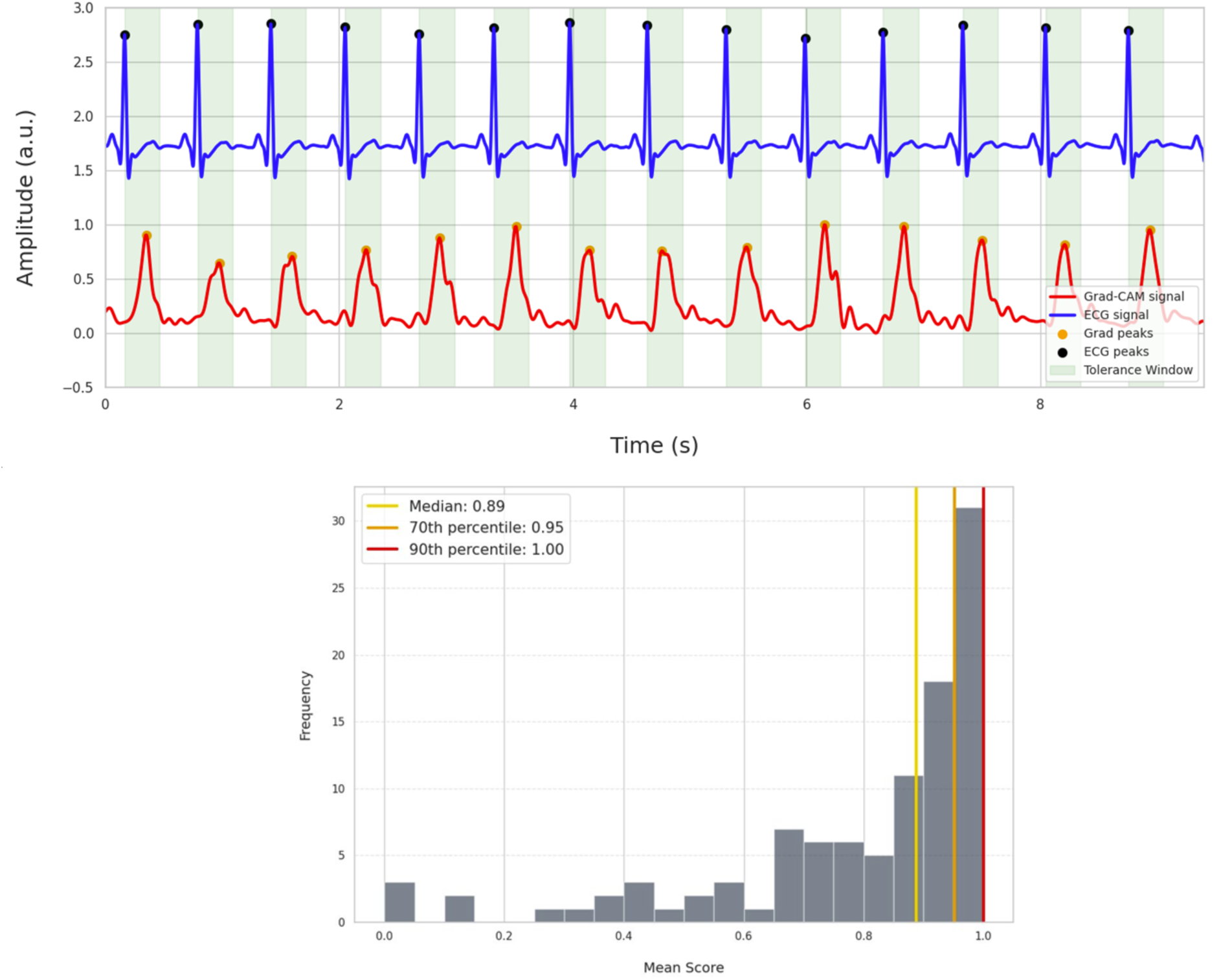
Temporal alignment and consistency of model-derived gradients with cardiac activity. **Top.** Temporal synchronisation between ECG R-peaks and gradient maxima. Representative example illustrating the alignment between detected ECG R-peaks and peak gradient intensities within a tolerance window of 0.3 s, with a slight temporal offset between the two signals. **Bottom.** Distribution of averaged temporal synchronisation scores per patient for the classification of Group 1 versus valvular heart disease (Group 3.[1–2–3]) during breath-hold acquisitions.

## Discussion

This study demonstrates the feasibility of contactless ultrasound chest vibration mapping in a clinical setting. It shows that the multipoint airborne ultrasound surface motion camera developed for this purpose, combined with deep learning algorithms, enables discrimination between healthy individuals and patients with respiratory or cardiac disease. The SMC-captured signals exhibit structured spatial and spectral organisation that can be exploited through data-driven models which, importantly, remain amenable to physiological interpretation.

### Thoracic vibration signatures separate health and disease

#### Respiratory patients

The ability of SMC to discriminate patients with respiratory disorders proved excellent. The AUC of the corresponding ROC curve was 0.90±0.07, hovering at the boundary between “excellent” and “outstanding” discrimination according to the classification proposed by Hosmer and Lemeshow ^22^. Discriminative performance peaked when comparing healthy individuals with patients classified as having severe respiratory disease. Yet performance in patients with the mildest of abnormalities, namely normal pulmonary function testing with abnormal chest computed tomography (our P1-1 group), was comparable to the overall estimate. In the literature, we could not find published data describing the performance of any diagnostic procedure to generically differentiate “respiratory normality” from “respiratory abnormality” irrespective of any pre-established diagnosis, whether based on clinical examination or on a technology-based approach. The closest reports we identified correspond to recent data-driven classification studies comparing multiple but pre-characterised respiratory conditions with healthy controls, based on analyses of expired CO2 dynamics ^23^ or of exhaled volatile organic compounds ^24,25^. These approaches achieved correct to excellent discriminative performance (e.g. AUC 0.74 [95% CI: 0.56-0.92] for CO2 dynamics ^23^; AUC 0.92 ±0.01 for volatile compound ^24^) but, unlike ours, did not follow a diagnostic-agnostic framework and were centred on predefined disease categories (including COPD, asthma, interstitial lung diseases, or pulmonary vascular diseases) or on curated datasets. In contrast, the medical literature abounds with studies assessing the performance of procedures targeting specific diagnoses, some of which align with our concept of an augmented clinical examination but in a focused manner (as in vibration response imaging derived from breath sound analysis for the detection of pleural effusions ^26^, or multichannel lung sound recording for the identification of lung fibrosis ^27^ or asthma ^28^). Such studies and procedures do not constitute directly relevant comparators to the present approach.

#### Cardiac patients

In contrast to the respiratory cohort, cardiac patients were included according to predefined diagnostic categories. In this setting, the AUCs of the ROC curves comparing healthy participants to different groupings of cardiac patients fell within the acceptable-to-excellent discrimination ranges ^22^. The AUC for the “healthy vs. all cardiac patients” comparison was 0.76 ± 0.10 during free-breathing acquisitions and 0.78 ± 0.08 during apnoea acquisitions, with a maximal value of 0.90 ± 0.10 when comparing healthy individuals to patients with aortic stenosis. These differences across groups may be partly explained by the acquisition geometry, better suited to capturing signals from the central aortic area than from the more lateral mitral region, which may have been only partially covered in some participants. Temporal variations in the signal related to heart rate, breathing pattern, and cardiac function^13,29^, together with interindividual differences in thoracic morphology, tissue composition, and posture ^29^, may also have contributed to signal heterogeneity. The higher performance observed in aortic stenosis may further reflect the relative homogeneity of this subgroup, in contrast to mitral valve disease, which encompasses more diverse pathophysiological mechanisms and associated thoracic vibration patterns. Notably, discriminative signals were also observed in cardiac subgroups without mitral regurgitation, indicating that detectable vibration patterns extend beyond valvular flow disturbances.

The cardiac SMC performance metrics suggest that it may offer advantages over clinical examination for the dichotomous identification of valvular heart disease. Indeed, a recent systematic review reported reasonable specificity but highly variable sensitivity (possibly as low as 30%) for clinical examination, with performance strongly dependent on the valve involved, the nature of the lesion (stenosis, regurgitation, or both), disease severity, and examiner expertise ^30^. At the other end of the spectrum, studies show that AI-assisted analysis of cardiac sounds can achieve very high accuracy, exceeding 95% ^31–33^. Such initiatives have been reported to increase the detection of previously unrecognised valvular disease, including in asymptomatic individuals ^34^, which represents a major unmet clinical need. These approaches, however, rely on contact-based acoustic recordings obtained under controlled conditions, with acquisition procedures that can be time-consuming and performance dependent on precise sensor positioning (as in the case of phonocardiography and cardiac seismography). For cardiomyopathies (Group 3.[4–5]), auscultatory detection is particularly challenging, and the role of digital auscultation remains less well established than for valvular disease. In this context, the SMC performance (AUC 0.74 ± 0.13 during free breathing) suggests potential value for the non-invasive assessment of this heterogeneous group of patients. Notably, cardiac performances may further improve with device optimisation.

### Model interpretation follows a structure–function framework

SMC offers major practical advantages, including contactless acquisition, no requirement for precise sensor positioning, and low-burden, operator-independent use. Its performance positions it as a potential extension of clinical examination, with potential to support the identification of abnormal pulmonary and cardiac states upstream of specialized procedures. However, the highly complex vibration patterns captured by SMC can only be meaningfully exploited through data-driven modelling, as is the case for other approaches relying on complex physiological signals ^23–25,27,28,31–33^. Model interpretability is central to the value of any AI-based diagnostic approach. Here, interpretability was explored using gradient-weighted class activation mapping (Grad-CAM), applied to the final convolutional layers of the models ^35^. This method generates spatially resolved maps highlighting the regions contributing most to model decisions, thereby providing access to the spatial organisation of the underlying vibration signals. In doing so, it moves beyond purely black-box prediction ^36,37^ and supports a degree of structure–function interpretation. Caution is warranted, however, as these representations, while informative, constitute indirect evidence and do not establish underlying physiological mechanisms ^38^

#### Respiratory models

In the respiratory modelling part of the study, the captured signals reflect how an input vibration is transformed into a distributed output pattern by the combined mechanical properties of the airways, lung parenchyma, pleural cavities, and chest wall. Because respiratory mechanics are well known to be frequency-dependent ^39–41^, SMC-derived vibratory maps should exhibit this property. In addition, the respiratory system is topographically inhomogeneous, with marked regional variations in its mechanical properties arising from both anatomical and physical factors. For example, the pyramidal form of the lungs implies that lung volume and lung expansion increase from the upper to the lower thorax ^42^, where, in addition, the presence of the heart involves right-to-left asymmetry. Moreover, the lungs are highly anisotropic, exhibiting a slinky-like gravity-dependent density gradient that contributes to regional variations in resistance and compliance ^43,44^. Together, these features imply that the response to a vibratory input should display spatial organisation in addition to its frequency dependence, and that the degree and nature of this frequency dependence should vary topographically. This has been observed previously using vocalisation as an endogenous and time-varying vibratory input ^14^, and the same principles are expected to apply to an exogenous and stable vibratory input such as that used in the present study, namely an oscillometry device ^45,46^. With this in mind, the admittance-based representations derived from our data, together with their decomposition into conductance and susceptance, appear highly compatible with physiological predictions. Indeed, in normal individuals, the observed maps exhibit non-random structured patterns that display consistent spatial organisation, with regional differences between upper and lower thoracic areas and between the two hemithoraces. In addition, their frequency dependence follows coherent trends, with progressive changes in both amplitude and spatial distribution across the explored frequency range. The respective behaviours of conductance and susceptance further reinforce this interpretation, as they exhibit distinct yet complementary spatial profiles, consistent with their association with dissipative and elastic components of the system ^46^. In contrast, in patients, the admittance maps depart from this organised behaviour and exhibit alterations in both their spatial structure and frequency dependence. The previously observed regional organisation becomes less consistent and less structured, with a tendency towards more heterogeneous and, in some cases, more spatially diffuse patterns. This is particularly apparent in some COPD cases (see illustrative examples in Results). The above changes are accompanied by modifications in the frequency-dependent behaviour of the signals, with altered amplitude profiles and less coherent spatial evolution across frequencies. The respective behaviours of conductance and susceptance are also affected, with a reduction in the complementarity observed in normal individuals, suggesting a disruption in the balance between dissipative and elastic components of the respiratory system ^41,46^. These features are compatible with disease altering the propagation of mechanical oscillations through the respiratory system, a phenomenon well documented in the physiological and clinical literature ^47,48^, including in patients without established lung function abnormalities ^49,50^, as in Group 2-1 of our study population. Taken together, these findings support the notion that the models used to discriminate healthy individuals from respiratory patients do not rely on arbitrary features, but rather capture physiologically meaningful signatures of how mechanical oscillations propagate through the respiratory system.

#### Cardiac models

In the cardiac modelling part of the study, the spatial distribution of gradients provides insight into the physiological basis of the decision-making process. Indeed, the regions identified by gradient analysis broadly coincide with classical cardiac auscultation areas ^51^, in particular the mitral and xiphoid regions, suggesting that spatial features consistent with established clinical landmarks are being leveraged. These observations provide a plausible mechanistic basis for the discriminative performance of the cardiac models. However, the areas of highest gradient intensity only partially overlap with the antenna coverage, which, with the prototype device used in this study, may have involved incomplete sampling of the mitral area in certain participants. Some physiologically informative regions are therefore not optimally captured with the current acquisition geometry, leaving room for performance improvement. The localisation of maximal gradients in inferolateral regions nevertheless appears somewhat counterintuitive in light of the observed discriminative performance, which was highest for aortic stenosis, a condition expected to predominantly involve more superior thoracic regions. This apparent discrepancy may reflect the fact that the recorded signals do not solely capture local mechanical activity but also include propagated vibration patterns. In this context, the models may be sensitive to seismic-like waves conveying information from cardiac structures beyond their point of origin, leading to informative signatures detectable at a distance from the primary source ^52^. The performance hierarchy according to disease classification also provides some physiology-based validation of the models. Indeed, the significantly higher discriminative performance observed for aortic stenosis may derive from the generation of high-velocity, turbulent transvalvular jets ^53,54^, known to produce substantial mechanical energy and to be conducive to vibration patterns that propagate at a distance, to the point of clinical detection beyond the thorax (e.g. neck vessels). In contrast, mitral conditions may give rise to more heterogeneous mechanical signatures, reflecting the variable geometry and direction of mitral jets ^54,55^, their partial dissipation within the left atrium ^55^, and the more posterior anatomical position of the mitral valve, which may limit efficient transmission to the anterior thoracic surface. Cardiac insufficiency, by comparison, is not characterised by focal high-velocity flow, but rather by more diffuse and lower-amplitude mechanical activity, which should result in less spatially coherent signals and reduced transmission to the measurement surface. Taken together, our observations, based on Grad-CAM topography and the hierarchy of discriminative performance across disease categories, suggest that the models capture spatially organised, physiologically meaningful patterns, while also highlighting the current limitations of the acquisition geometry and the need for cautious interpretation. They further point to a potential avenue for optimisation, whereby improved antenna positioning or expanded coverage could enhance the capture of relevant mechanical signals and, consequently, model performance.

### Study limitations and strengths

Several limitations of the present study should be acknowledged. First, the use of a pilot device, the relatively limited sample size leading to some small subgroups, the age impbalance between the healthy participants and the patients, and the absence of certain clinically relevant conditions in the case mix constrain the robustness of the findings and their generalisability. For example, no pleural disorders were represented in the respiratory subset, although their pronounced effects on thoracic vibration transmission would make them particularly informative test cases. Likewise, no complex valvular conditions were included in the cardiac subset. Second, the grouping strategy adopted for the cardiac cohort (participants selected on predefined conditions and severity levels) reflects the constraints of a pilot design and should not be taken as a definitive representation of the broader cardiac population. Addressing this limitation will require real-world studies including patients with suspected cardiac disorders who are not selected a priori on the basis of a predefined diagnosis. Third, several technical and operational limitations related to the use of a research prototype device must be considered. Limited thoracic coverage precluded homogeneous acquisitions over the cardiac auscultation areas, as discussed above, and constrained adaptation to individual anatomical configurations. Resorting to a oscillometry device, instead of vocalisations as initially described ^14^, for stimulus standardisation purposes introduces additional procedural complexity. In addition, the feasibility data highlight practical constraints inherent to the acquisition chain, with failures arising from inability to perform oscillometry measurements, oscillometry–SMC synchronisation issues, and suboptimal signal-to-noise ratios. Real-time visualisation of the acquired signal was not available, increasing the likelihood of post hoc rejection of certain acquisitions. Taken together, these constraints suggest that the results of the present study should be regarded as conservative estimates, with room for improvement through technical refinement. More generally, signal acquisition may be affected by inter-individual variability in morphology, posture, or movement, including conditions such as advanced age or neurological disorders, which could alter thoracic mechanics and introduce variability in the measurements. These factors warrant specific investigation in future studies. Fourth, different algorithms were used for respiratory and cardiac classification tasks, trained on datasets of differing size and representativeness. As with any data-driven approach, the stability and generalisability of the models will require confirmation in larger datasets and across different acquisition conditions. At this stage, this also precluded any meaningful attempt to directly discriminate between respiratory and cardiac patients. Fifth, although the interpretability analyses provide valuable insights into the spatial organisation of the signals used by the models, they remain indirect and should be interpreted with caution. The observed structure–function relationships are inferred from model behaviour and do not constitute direct evidence of underlying physiological mechanisms. Further work will be required to better characterise the physical and biological bases of the recorded vibration patterns.

Despite these limitations, the study also has several strengths. It introduces a novel, disruptive approach to the assessment of cardiopulmonary mechanics, capable of capturing spatially resolved vibration patterns in a contactless, low-burden, and operator-independent manner. This is achieved with rapid acquisition times (on the order of a few tens of seconds) and minimal requirements for patient cooperation, which proved true even in elderly and frail individuals with severe pulmonary or cardiac diseases. The technique is risk-free and does not involve exposure to ionising radiation. The study design, based on prospective acquisitions in clinically characterised participants without extensive experimental constraints, approaches real-world conditions and supports the practical relevance of the findings. In this regard, the diagnostic-agnostic framework adopted in the respiratory component of the study represents a departure from conventional approaches focused on predefined disease categories. More broadly, this approach distinguishes itself from existing approaches by combining contactless acquisition, multipoint spatial mapping, and data-driven analysis within a unified framework. In addition, the combination of data-driven modelling with physiologically interpretable features provides a promising avenue for bridging computational analysis and clinical reasoning.

### Perspectives and conclusions

Next steps involve determining whether SMC can support practical diagnostic pathways, for example by helping to discriminate between cardiac and respiratory causes of acute dyspnoea in emergency or primary care settings, including in patients with mixed or ambiguous presentations such as encountered in cardiopulmonary overlap conditions. It will also be important to assess whether the approach can operate at a finer level of granularity, enabling the identification of specific conditions (e.g. pleural disorders) or supporting longitudinal monitoring. SMC could also prove useful for documenting disease course, detecting exacerbations, and assessing treatment effects.

In conclusion, our findings indicate that thoracic vibration transmission patterns contain spatially and spectrally organised information that can be harnessed for clinical purposes through data-driven modelling. The computational models used in this study proved amenable to physiological interpretation, which is critical for clinical relevance. The SMC represents a novel, quantitative and objectivised extension of long-standing clinical examination, whereby physicians have historically relied on palpation of the chest wall to appreciate vibration-related phenomena such as fremitus or thrills. Our results support positioning SMC as a putative immediate extension of clinical examination, operating upstream of imaging and potentially contributing to the selection of subsequent diagnostic investigations.

## Data Availability

Material availability
1. Data
All individual participant data collected during the study, duly deidentified, will be made available immediately following publication, with no end date. Access will be granted to researchers submitting a methodologically sound proposal approved by an independent review committee (learned intermediary) designated for this purpose. Data will be shared exclusively for the purpose of achieving the objectives outlined in the approved proposal. Data requests should be directed to the corresponding author. Requestors will be required to sign a data access agreement as a condition of access.
2. Code
The code used to generate the results reported in this study will be made available upon acceptance of the manuscript, under the same provisions as those governing data availability.

## Acknowledgements

The authors are grateful to the Clinical Research Department, Caen university hospital for their methodological and logistical support; to Ms Laurence Lamoureux and Claire Lemperière, research nurses, for their help in acquiring the data; and to Frédéric Giral and Guillaume Knispel for support and advice.

## Declarations

### Authors contributions (CredIT taxonomy)

1. Conceptualization: TS, MC, ES, AM, RKI
2. Methodology: MS, LD, MC, AZ, FW, ES, RKI
3. Formal analysis: MS, LD, FW
4. Investigation: MC, ES, AM, AH, FW
5. Data Curation: FW, LD
6. Data Interprestation: TS, ES, MS, LD
7. Writing - Original Draft: TS, LD, MC,ES
8. Writing - Review & Editing: TS, MS, FW, MF, RKI
9. Supervision: PMF
10. Project administration: PMF
11. Accountability: all authors

## Material availability

### 1. Data

All individual participant data collected during the study, duly deidentified, will be made available immediately following publication, with no end date. Access will be granted to researchers submitting a methodologically sound proposal approved by an independent review committee (“learned intermediary”) designated for this purpose. Data will be shared exclusively for the purpose of achieving the objectives outlined in the approved proposal. Data requests should be directed to the corresponding author. Requestors will be required to sign a data access agreement as a condition of access.

### 2. Code

The code used to generate the results reported in this study will be made available upon acceptance of the manuscript, under the same provisions as those governing data availability.

## Use of generative intelligence in writing the manuscript

No part of this manuscript has been produced using generative artificial intelligence.

## Electronic supplement

**Electronic supplement to**

Contactless ultrasound chest vibration mapping discriminates respiratory and cardiac patients from healthy individuals.

Eric SALOUX, Loïc DEMORE, et al.

## Supplementary tables

**Table S1.**
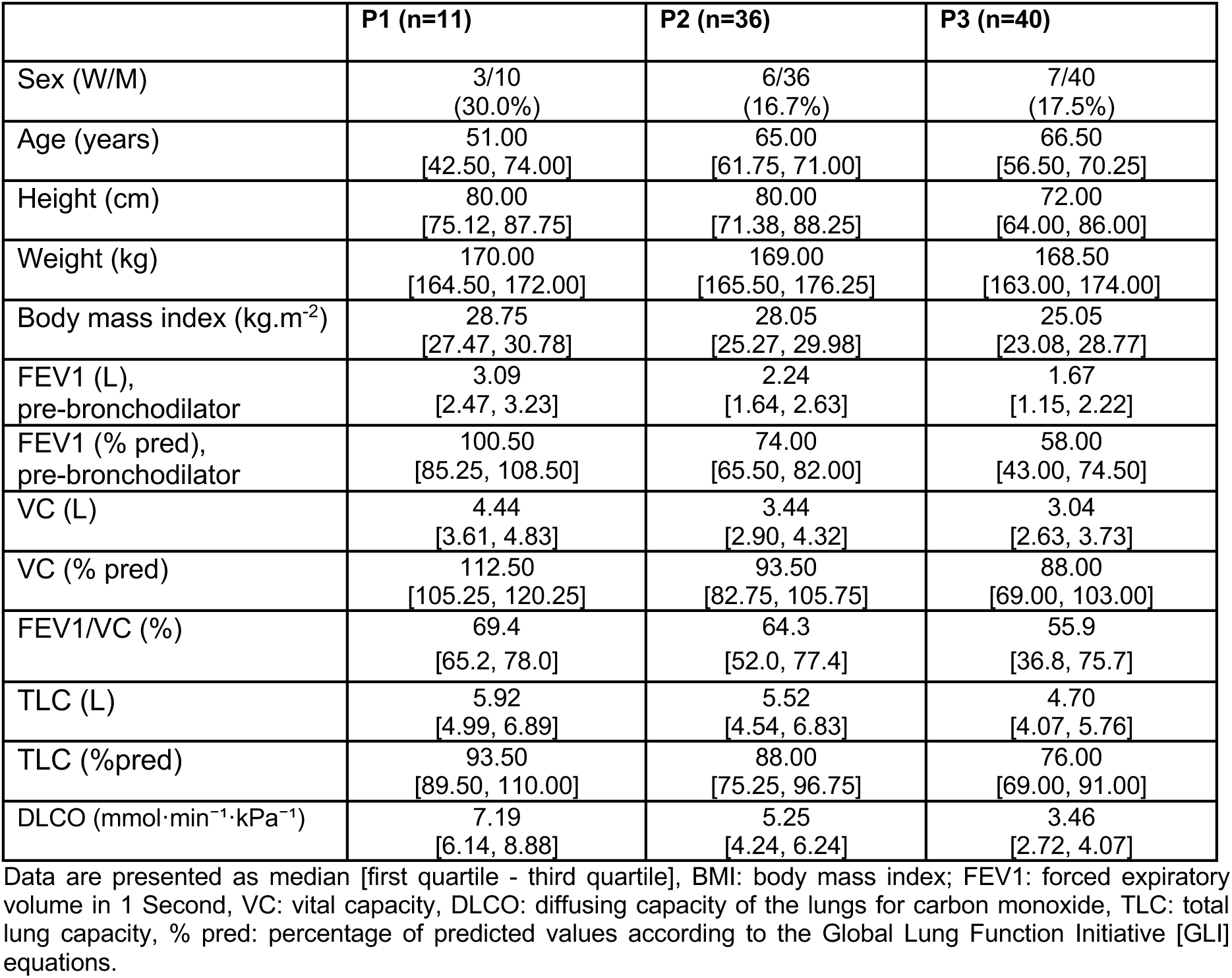
Characteristics of the respiratory patients retained for analysis (n = 87) according to subgroup classification.

**Table S2.**
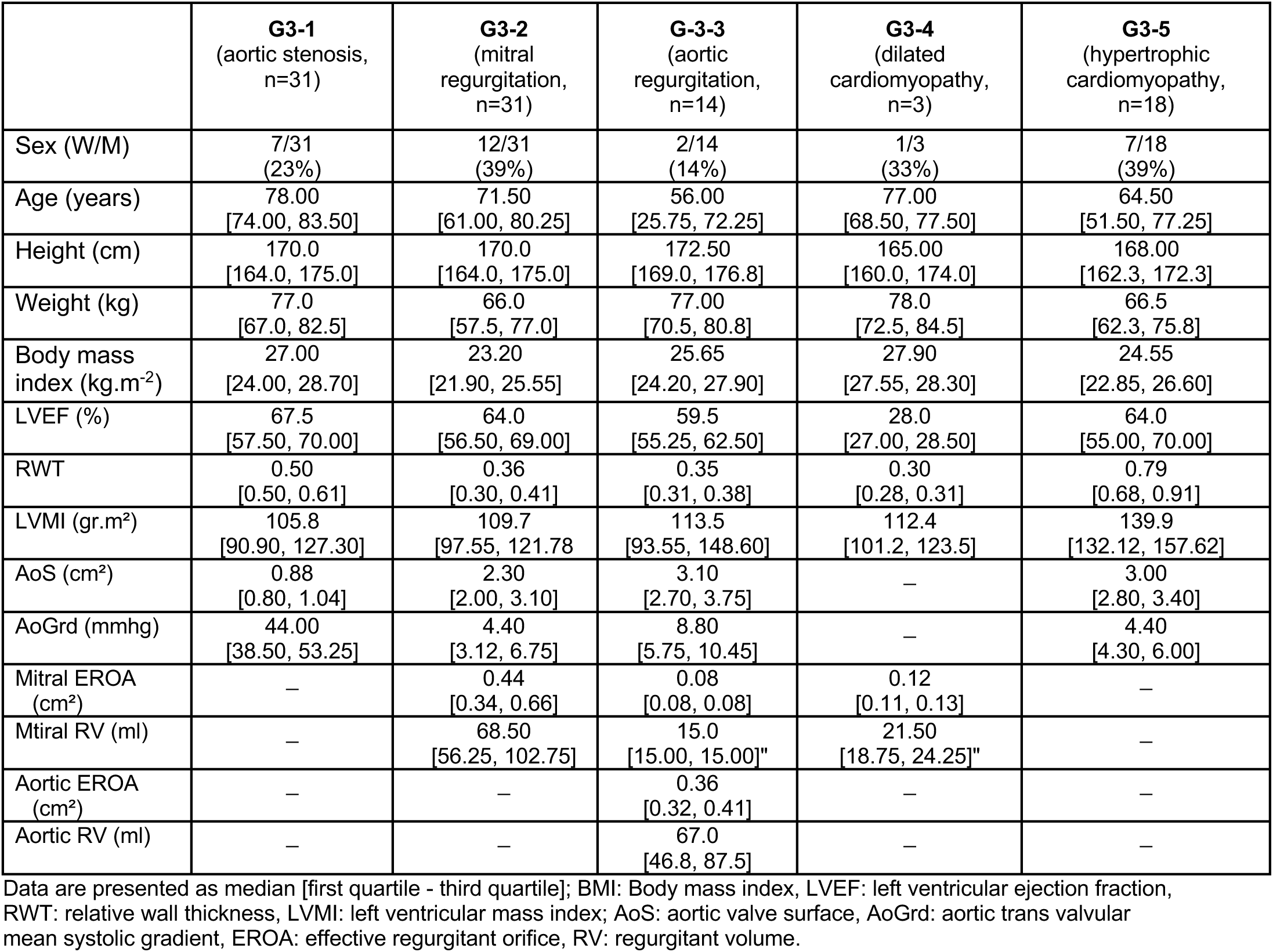
Characteristics of the cardiac patients retained for analysis (n = 97).

**Table S3.**
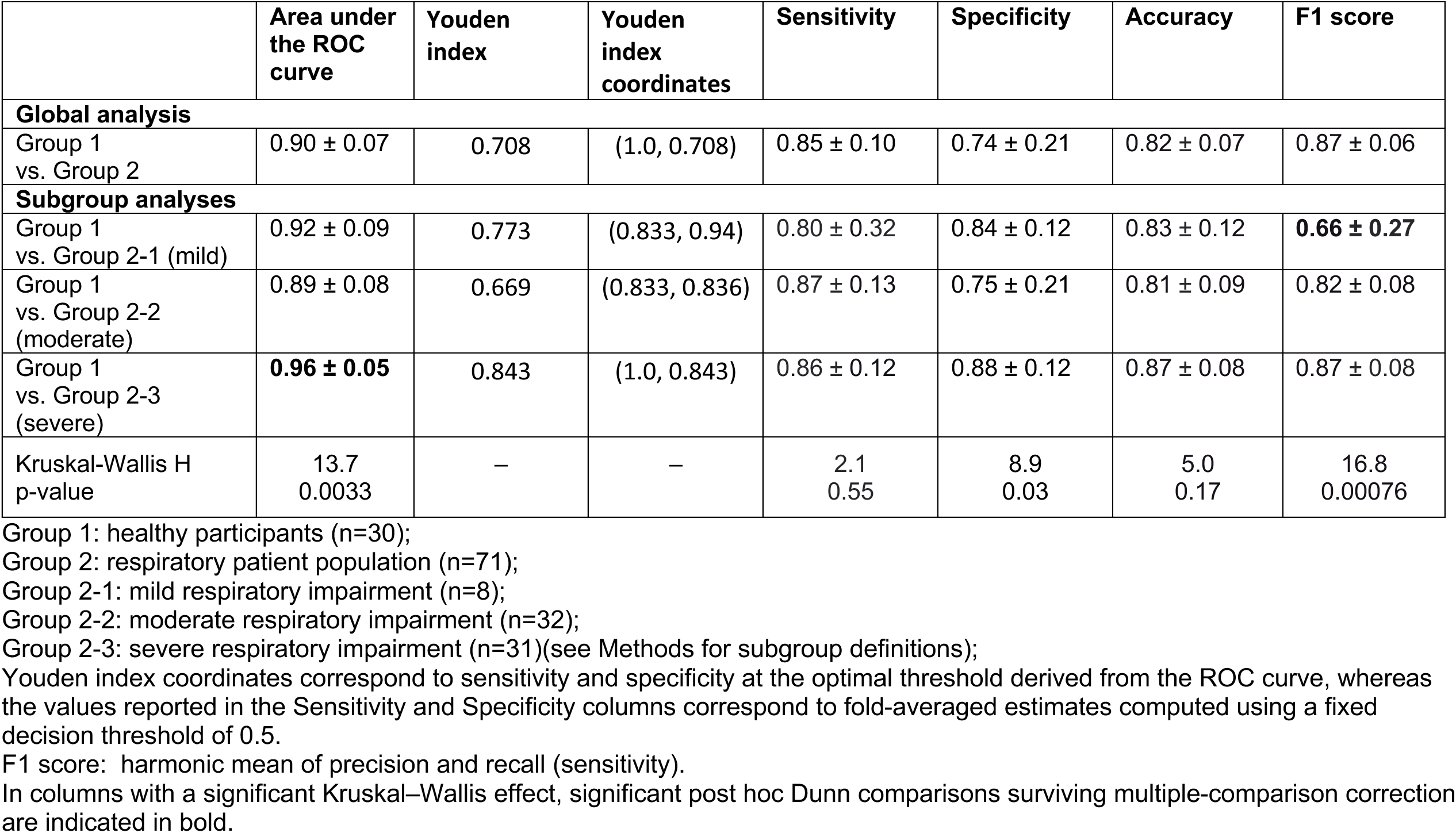
Performance metrics of the surface motion camera for discriminating healthy participants from respiratory patients.

**Table S4.**
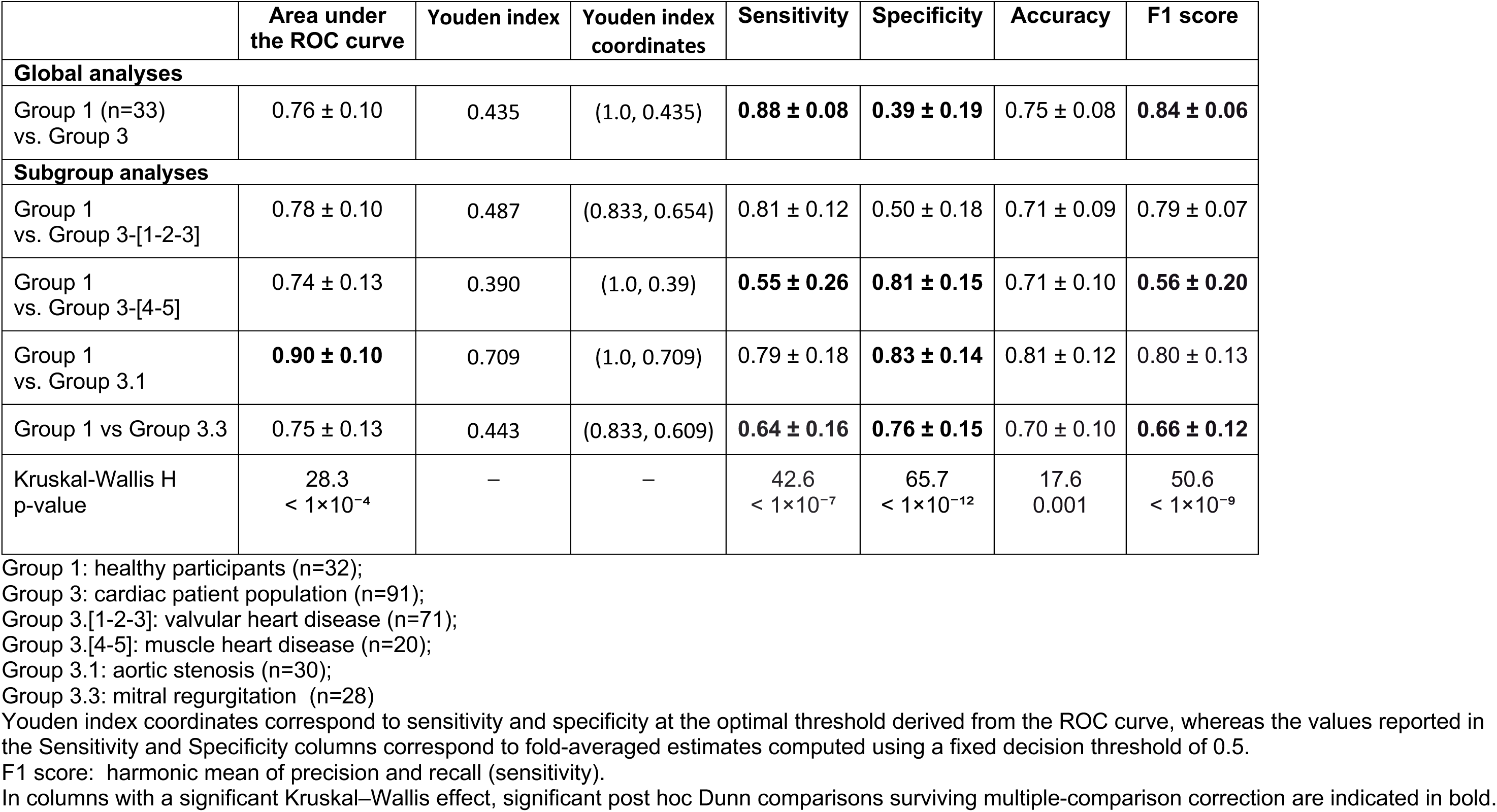
Performance metrics of the surface motion camera for discriminating healthy participants from cardiac patients (acquisitions performed during free breathing).

**Table S5.**
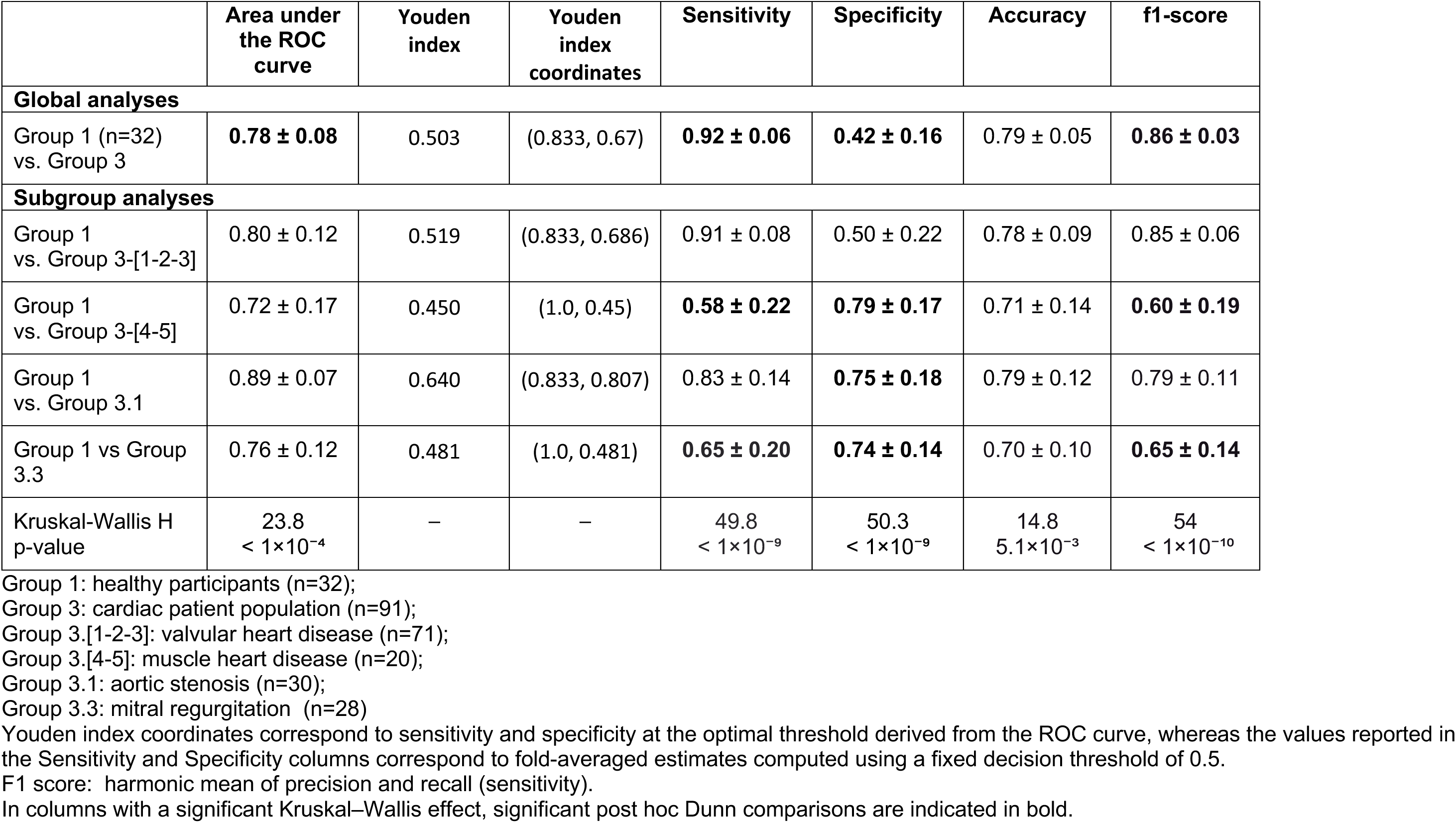
Performance metrics of the surface motion camera for discriminating healthy participants from cardiac patients (acquisitions performed during breath-holding).

**Table S6.**
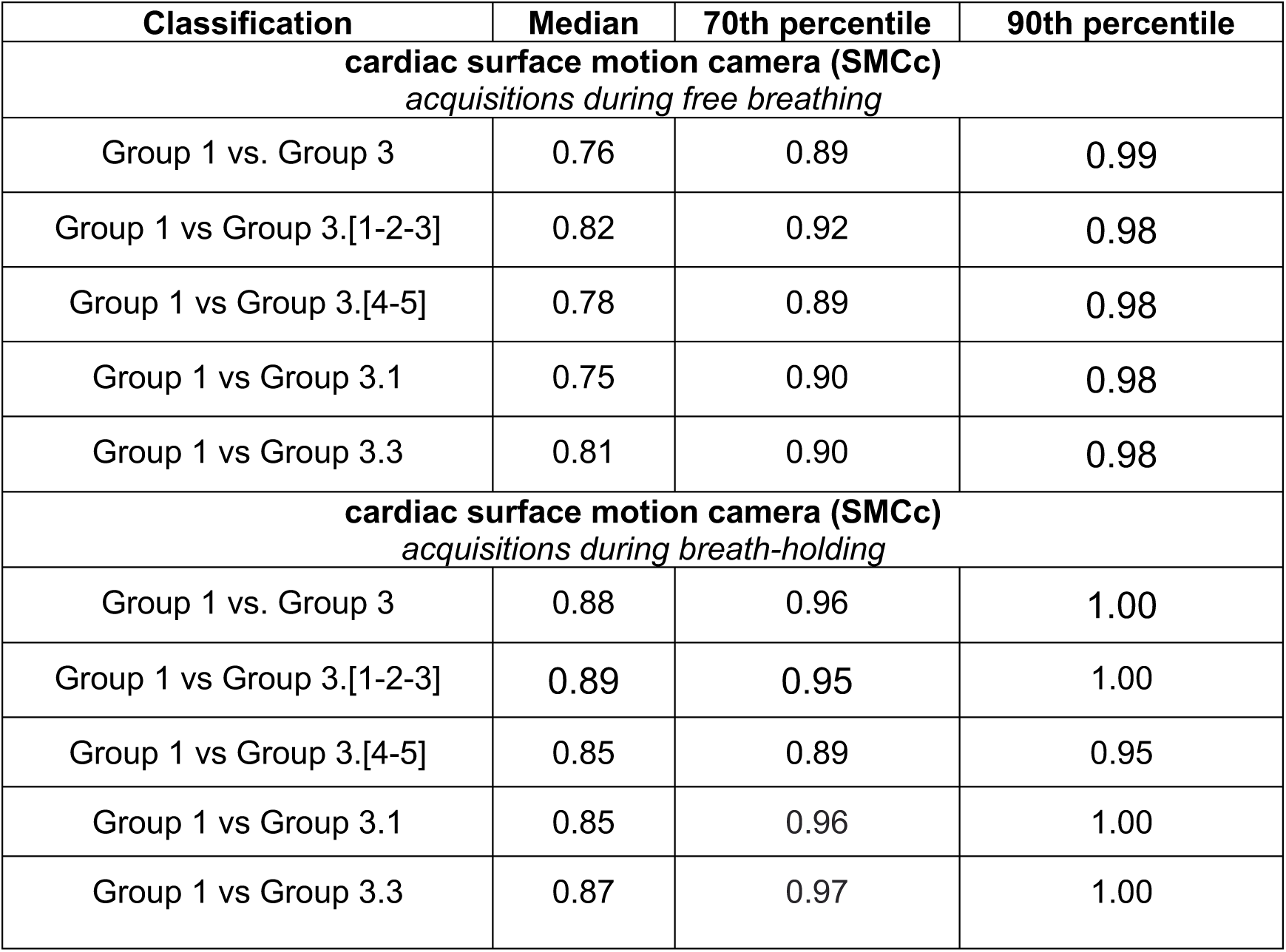
Summary of temporal synchronisation scores between gradient maxima and ECG signals across classification tasks and acquisition conditions.

## Supplementary Figures

**Figure S1.**
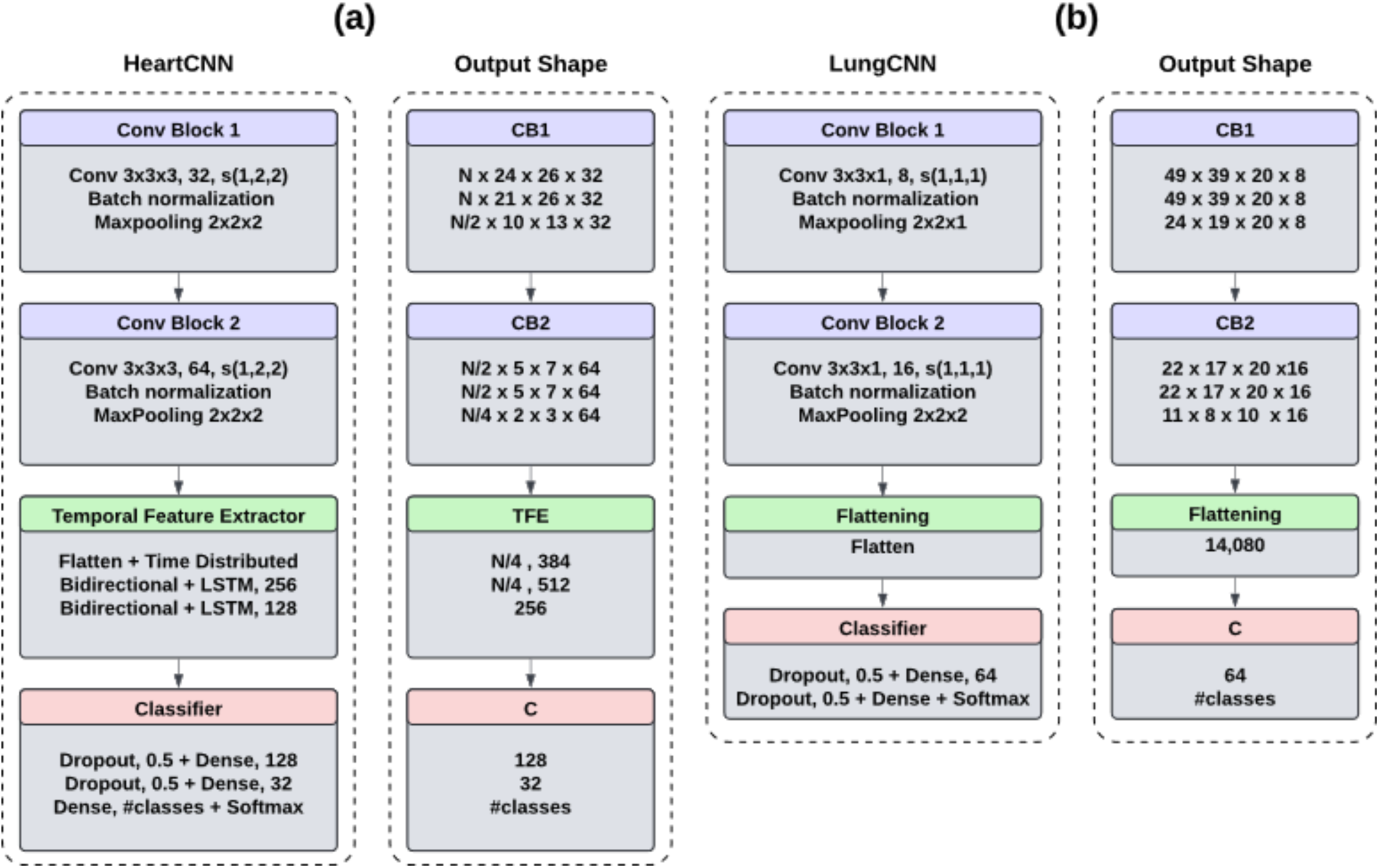
Schematic representation of the proposed network architectures. **A.** cardiac convolutional neural network; **B.** pulmonafry convolutional neural network. N denotes the temporal dimension of the surface moction camera cardiac acquisitions (SMCc); CB, convolutional block; TFE, temporal feature extractor; C, classifier; and LSTM, long short-term memory.

## Supplementary videos

[as of April 27 2026, the supplementary videos can be downloaded at this temporary address: http://urlr.me/uEk6Nr]

**Video S1.** Respiratory-related thoracic dynamic surface vibration map in a healthy participant (Group 1). The left panel (“spatial slice”) shows the spatial admittance map at the frequency F. On the right, the top panel (“frequential slice (F,X)”) shows the frequential slice of admittance map along Y axis, the middle panel (“frequential slice (F,Y)”) shows the frequential slice of admittance map along X axis, the bottom panel (“frequential signal along” shows the frequential signal along (X,Y) pixel).

**Video S2.** Cardiac-related thoracic dynamic surface vibration map in a healthy participant (Group 1). The left panel (“spatial slice”) shows the spatial seismocardiogram map at time T. On the right, the top panel (“temporal slice(T,X)”) shows the temporal slice of seismocardiogram map along Y axis, the middle panel (“temporal slice (T,Y)”) shows the temporal slice of seismocardiogram map along X axis, the bottom panel (“temporal signal along” shows the seismocardiogram signal along (X,Y) pixel.

**Video S3**. Respiratory-related thoracic dynamic surface vibration map in a participant with severe lung function abnormalities (Group 2-3). The left panel (“spatial slice”) shows the spatial admittance map at the frequency F. On the right, the top panel (“frequential slice(F,X)”) shows the frequential slice of admittance map along Y axis, the middle panel (“frequential slice (F,Y)”) shows the frequential slice of admittance map along X axis, the bottom panel (“frequential signal along” shows the frequential signal along (X,Y) pixel).

**Video S4.** Cardiac-related thoracic surface dynamic vibration map in a participant with aortic stenosis (Group 3-1). The left panel (“spatial slice”) shows the spatial seismocardiogram map at time T. On the right, the top panel (“temporal slice(T,X)”) shows the temporal slice of seismocardiogram map along Y axis, the middle panel (“temporal slice (T,Y)”) shows the temporal slice of seismocardiogram map along X axis, the bottom panel (“temporal signal along” shows the seismocardiogram signal along (X,Y) pixel).

**Video S5.** Cardiac-related thoracic surface dynamic vibration map in a participant with restrictive cardiomyopathy (Group 3-5). The left panel (“spatial slice”) shows the spatial seismocardiogram map at time T. On the right, the top panel (“temporal slice(T,X)”) shows the temporal slice of seismocardiogram map along Y axis, the middle panel (“temporal slice (T,Y)”) shows the temporal slice of seismocardiogram map along X axis, the bottom panel (“temporal signal along” shows the seismocardiogram signal along (X,Y) pixel).

## References

1 Hippocrates. Hippocratic writings (translation by E.T. Withington, 1927-1931; Loeb Classical Library. Cambridge, MA, USA: Harvard University Press). (circa 400 B.C).

2 Laennec, R. De l’auscultation médiate., (J.-A Brosson & J.-S Chaudé, 1819).

3 Association of American Medical Colleges. Core entrustable professional activities for entering residency: curriculum developers’ guide. (Association of American Medical Colleges, 2014).

4 European Respiratory Society. Harmonised Education in Respiratory Medicine for European Specialists (HERMES): adult respiratory medicine syllabus. (European Respiratory Society, 2017).

5 General Medical Council. Outcomes for graduates., (General Medical Council, 2020).

6 Bickley, L. & Szilagyi, P. Bates’ guide to physical examination and history taking, 14th edition. (Wolters Kluwer, 2024).

7 Maitre, B., Similowski, T. & Derenne, J. P. Physical examination of the adult patient with respiratory diseases: inspection and palpation. Eur Respir J 8, 1584–1593 (1995).

8 Smith-Bindman, R. et al. Projected Lifetime Cancer Risks From Current Computed Tomography Imaging. JAMA Internal Medicine 185, 710–719 (2025). 10.1001/jamainternmed.2025.0505

9 Kwee, R. M., Toxopeus, R. & Kwee, T. C. Imaging overuse in the emergency department: The view of radiologists and emergency physicians. European Journal of Radiology 176, 111536 (2024). 10.1016/j.ejrad.2024.111536

10 Khalili, A., Drummond, J., Ramjattan, N., Zeltser, R. & Makaryus, A. N. Diagnostic and treatment utility of echocardiography in the management of the cardiac patient. World J Cardiol 12, 262–268 (2020). 10.4330/wjc.v12.i6.262

11 Nierat, M. C. et al. Physiological Validation of an Airborne Ultrasound Based Surface Motion Camera for a Contactless Characterization of Breathing Pattern in Humans. Front Physiol 10, 680 (2019). 10.3389/fphys.2019.00680

12 Shirkovskiy, P. et al. Airborne ultrasound surface motion camera: Application to seismocardiography. Applied Physics Letters 112 (2018). 10.1063/1.5028348

13 Sadhukhan, D. et al. Non-Contact Assessment of Cardiac Velocity Profiles Using Ultrasound Based Surface Motion Camera: A Feasibility Study. IEEE Journal of Biomedical and Health Informatics 30, 586–595 (2026). 10.1109/JBHI.2025.3577130

14 Wintzenrieth, F. et al. Airborne ultrasound for the contactless mapping of surface thoracic vibrations during human vocalizations: A pilot study. AIP Advances 14 (2024). 10.1063/5.0187945

15 Hall, G. L. et al. Official ERS technical standard: Global Lung Function Initiative reference values for static lung volumes in individuals of European ancestry. Eur Respir J 57 (2021). 10.1183/13993003.00289-2020

16 Quanjer, P. H. et al. Multi-ethnic reference values for spirometry for the 3-95-yr age range: the global lung function 2012 equations. Eur Respir J 40, 1324–1343 (2012). 10.1183/09031936.00080312

17 Stanojevic, S. et al. Official ERS technical standards: Global Lung Function Initiative reference values for the carbon monoxide transfer factor for Caucasians. Eur Respir J 50 (2017). 10.1183/13993003.00010-2017

18 Lang, R. M. et al. Recommendations for cardiac chamber quantification by echocardiography in adults: an update from the American Society of Echocardiography and the European Association of Cardiovascular Imaging. J Am Soc Echocardiogr 28, 1–39.e14 (2015). 10.1016/j.echo.2014.10.003

19 Vahanian, A. et al. 2021 ESC/EACTS Guidelines for the management of valvular heart disease. Eur Heart J 43, 561–632 (2022). 10.1093/eurheartj/ehab395

20 Zoghbi, W. A. et al. Recommendations for Noninvasive Evaluation of Native Valvular Regurgitation: A Report from the American Society of Echocardiography Developed in Collaboration with the Society for Cardiovascular Magnetic Resonance. J Am Soc Echocardiogr 30, 303–371 (2017). 10.1016/j.echo.2017.01.007

21 Savitzky, A. & Golay, M. J. E. Smoothing and Differentiation of Data by Simplified Least Squares Procedures. Analytical Chemistry 36, 1627–1639 (1964). 10.1021/ac60214a047

22 Hosmer, D. & Lemeshow, S. Applied logistic regression, 2nd edition, pp 156-164. 156–164 (Wiley, 2000).

23 John, T. et al. Detection of structural pulmonary changes with real-time high-fidelity analysis of expiratory CO(2). J Breath Res 19 (2025). 10.1088/1752-7163/adf253

24 Tian, J. et al. Exhaled volatile organic compounds as novel biomarkers for early detection of COPD, asthma, and PRISm: a cross-sectional study. Respir Res 26, 173 (2025). 10.1186/s12931-025-03242-5

25 Yu, K. L. et al. Exhaled Breath Analysis Using a Novel Electronic Nose for Different Respiratory Disease Entities. Lung 203, 14 (2025). 10.1007/s00408-024-00776-1

26 Anantham, D., Herth, F. J., Majid, A., Michaud, G. & Ernst, A. Vibration response imaging in the detection of pleural effusions: a feasibility study. Respiration 77, 166–172 (2009). 10.1159/000168784

27 Messner, E. et al. Multi-channel lung sound classification with convolutional recurrent neural networks. Comput Biol Med 122, 103831 (2020). 10.1016/j.compbiomed.2020.103831

28 Islam, M. A., Bandyopadhyaya, I., Bhattacharyya, P. & Saha, G. Multichannel lung sound analysis for asthma detection. Comput Methods Programs Biomed 159, 111–123 (2018). 10.1016/j.cmpb.2018.03.002

29 Taebi, A., Solar, B. E., Bomar, A. J., Sandler, R. H. & Mansy, H. A. Recent Advances in Seismocardiography. Vibration 2, 64–86 (2019). 10.3390/vibration2010005

30 Davidsen, A. H. et al. Diagnostic accuracy of heart auscultation for detecting valve disease: a systematic review. BMJ Open 13, e068121 (2023). 10.1136/bmjopen-2022-068121

31 Ding, S. J. et al. A Computer-Aided Heart Valve Disease Diagnosis System Based on Machine Learning. J Healthc Eng 2023, 7382316 (2023). 10.1155/2023/7382316

32 Popat, A. et al. Diagnostic Accuracy of AI Algorithms in Aortic Stenosis Screening: A Systematic Review and Meta-Analysis. Clin Med Res 22, 145–155 (2024). 10.3121/cmr.2024.1934

33 Thompson, W. R., Reinisch, A. J., Unterberger, M. J. & Schriefl, A. J. Artificial Intelligence-Assisted Auscultation of Heart Murmurs: Validation by Virtual Clinical Trial. Pediatr Cardiol 40, 623–629 (2019). 10.1007/s00246-018-2036-z

34 Martin, J., de Verteuil, J., Schriefl, A. & Denham, S. Increasing detection of heart valve disease in Farnborough Primary Care Network using auscultation artificial intelligence in the community pharmacy setting. International Journal of Pharmacy Practice 31, ii32-ii33 (2023). 10.1093/ijpp/riad074.039

35 Selvaraju, R. R. et al. Grad-CAM: Visual Explanations from Deep Networks via Gradient-Based Localization. Int. J. Comput. Vision 128, 336–359 (2020). 10.1007/s11263-019-01228-7

36 Rudin, C. Stop explaining black box machine learning models for high stakes decisions and use interpretable models instead. Nature Machine Intelligence 1, 206–215 (2019). 10.1038/s42256-019-0048-x

37 Tjoa, E. & Guan, C. A Survey on Explainable Artificial Intelligence (XAI): Toward Medical XAI. IEEE Trans Neural Netw Learn Syst 32, 4793–4813 (2021). 10.1109/tnnls.2020.3027314

38 Adebayo, J. et al. in Proceedings of the 32nd International Conference on Neural Information Processing Systems 9525–9536 (Curran Associates Inc., Montréal, Canada, 2018).

39 Fredberg, J. J. & Stamenovic, D. On the imperfect elasticity of lung tissue. J Appl Physiol (1985) 67, 2408–2419 (1989). 10.1152/jappl.1989.67.6.2408

40 Hantos, Z., Daróczy, B., Suki, B., Nagy, S. & Fredberg, J. J. Input impedance and peripheral inhomogeneity of dog lungs. J Appl Physiol (1985) 72, 168–178 (1992). 10.1152/jappl.1992.72.1.168

41 Suki, B. & Bates, J. H. Lung tissue mechanics as an emergent phenomenon. J Appl Physiol (1985) 110, 1111–1118 (2011). 10.1152/japplphysiol.01244.2010

42 Milic-Emili, J., Henderson, J. A., Dolovich, M. B., Trop, D. & Kaneko, K. Regional distribution of inspired gas in the lung. J Appl Physiol 21, 749–759 (1966). 10.1152/jappl.1966.21.3.749

43 Mead, J., Takishima, T. & Leith, D. Stress distribution in lungs: a model of pulmonary elasticity. J Appl Physiol 28, 596–608 (1970). 10.1152/jappl.1970.28.5.596

44 Komlosi, P. et al. Regional anisotropy of airspace orientation in the lung as assessed with hyperpolarized helium-3 diffusion MRI. J Magn Reson Imaging 42, 1777–1782 (2015). 10.1002/jmri.24950

45 Michaelson, E. D., Grassman, E. D. & Peters, W. R. Pulmonary mechanics by spectral analysis of forced random noise. J Clin Invest 56, 1210–1230 (1975). 10.1172/jci108198

46 Bates, J. H. T., Irvin, C. G., Farré, R. & Hantos, Z. Oscillation Mechanics of the Respiratory System. Comprehensive Physiology 1, 1233–1272 (2011). 10.1002/j.2040-4603.2011.tb00366.x

47 Oostveen, E. et al. The forced oscillation technique in clinical practice: methodology, recommendations and future developments. Eur Respir J 22, 1026–1041 (2003). 10.1183/09031936.03.00089403

48 Bednarek, M., Grabicki, M., Piorunek, T. & Batura-Gabryel, H. “Current place of impulse oscillometry in the assessment of pulmonary diseases.”. Respir Med 170, 105952 (2020). 10.1016/j.rmed.2020.105952

49 Berger, K. I. et al. Distal airway dysfunction identifies pulmonary inflammation in asymptomatic smokers. ERJ Open Res 2 (2016). 10.1183/23120541.00066-2016

50 Bhattarai, P. et al. Early detection of small airway dysfunction in smokers and people with COPD via forced oscillation technique and its association with biomarkers: a pilot study. Am J Physiol Lung Cell Mol Physiol 330, L211–l221 (2026). 10.1152/ajplung.00155.2025

51 Leatham, A. Auscultation of the heart. Lancet 2, 703–708 contd (1958). 10.1016/s0140-6736(58)91329-1

52 Inan, O. T. et al. Ballistocardiography and seismocardiography: a review of recent advances. IEEE J Biomed Health Inform 19, 1414–1427 (2015). 10.1109/jbhi.2014.2361732

53 Sabbah, H. N. & Stein, P. D. Turbulent blood flow in humans: its primary role in the production of ejection murmurs. Circ Res 38, 513–525 (1976). 10.1161/01.res.38.6.513

54 Yoganathan, A. P., He, Z. & Casey Jones, S. Fluid mechanics of heart valves. Annu Rev Biomed Eng 6, 331–362 (2004). 10.1146/annurev.bioeng.6.040803.140111

55 Yoran, C. et al. Dynamic aspects of acute mitral regurgitation: effects of ventricular volume, pressure and contractility on the effective regurgitant orifice area. Circulation 60, 170–176 (1979). 10.1161/01.cir.60.1.170

