## Supplementary material for "Contactless ultrasound chest vibration mapping discriminates respiratory and cardiac patients from healthy individuals"

### Electronic supplement

#### Supplementary tables

**Table S1. Characteristics of the respiratory patients retained for analysis (n = 87) according to subgroup classification.**

| | P1 (n=11) | P2 (n=36) | P3 (n=40) |
|---|---|---|---|
| Sex (W/M) | 3/10 (30.0%) | 6/36 (16.7%) | 7/40 (17.5%) |
| Age (years) | 51.00 [42.50, 74.00] | 65.00 [61.75, 71.00] | 66.50 [56.50, 70.25] |
| Height (cm) | 80.00 [75.12, 87.75] | 80.00 [71.38, 88.25] | 72.00 [64.00, 86.00] |
| Weight (kg) | 170.00 [164.50, 172.00] | 169.00 [165.50, 176.25] | 168.50 [163.00, 174.00] |
| Body mass index ($kg.m^{-2}$) | 28.75 [27.47, 30.78] | 28.05 [25.27, 29.98] | 25.05 [23.08, 28.77] |
| FEV1 (L), pre-bronchodilator | 3.09 [2.47, 3.23] | 2.24 [1.64, 2.63] | 1.67 [1.15, 2.22] |
| FEV1 (% pred), pre-bronchodilator | 100.50 [85.25, 108.50] | 74.00 [65.50, 82.00] | 58.00 [43.00, 74.50] |
| VC (L) | 4.44 [3.61, 4.83] | 3.44 [2.90, 4.32] | 3.04 [2.63, 3.73] |
| VC (% pred) | 112.50 [105.25, 120.25] | 93.50 [82.75, 105.75] | 88.00 [69.00, 103.00] |
| FEV1/VC (%) | 69.4 [65.2, 78.0] | 64.3 [52.0, 77.4] | 55.9 [36.8, 75.7] |
| TLC (L) | 5.92 [4.99, 6.89] | 5.52 [4.54, 6.83] | 4.70 [4.07, 5.76] |
| TLC (%pred) | 93.50 [89.50, 110.00] | 88.00 [75.25, 96.75] | 76.00 [69.00, 91.00] |
| DLCO ($mmol \cdot min^{-1} \cdot kPa^{-1}$) | 7.19 [6.14, 8.88] | 5.25 [4.24, 6.24] | 3.46 [2.72, 4.07] |

Data are presented as median [first quartile - third quartile], BMI: body mass index; FEV1: forced expiratory volume in 1 Second, VC: vital capacity, DLCO: diffusing capacity of the lungs for carbon monoxide, TLC: total lung capacity, % pred: percentage of predicted values according to the Global Lung Function Initiative [GLI] equations.

**Table S2. Characteristics of the cardiac patients retained for analysis (n = 97).**

| | **G3-1** (aortic stenosis, n=31) | **G3-2** (mitral regurgitation, n=31) | **G-3-3** (aortic regurgitation, n=14) | **G3-4** (dilated cardiomyopathy, n=3) | **G3-5** (hypertrophic cardiomyopathy, n=18) |
|---|---|---|---|---|---|
| Sex (W/M) | 7/31 (23%) | 12/31 (39%) | 2/14 (14%) | 1/3 (33%) | 7/18 (39%) |
| Age (years) | 78.00 [74.00, 83.50] | 71.50 [61.00, 80.25] | 56.00 [25.75, 72.25] | 77.00 [68.50, 77.50] | 64.50 [51.50, 77.25] |
| Height (cm) | 170.0 [164.0, 175.0] | 170.0 [164.0, 175.0] | 172.50 [169.0, 176.8] | 165.00 [160.0, 174.0] | 168.00 [162.3, 172.3] |
| Weight (kg) | 77.0 [67.0, 82.5] | 66.0 [57.5, 77.0] | 77.00 [70.5, 80.8] | 78.0 [72.5, 84.5] | 66.5 [62.3, 75.8] |
| Body mass index (kg.m$^{-2}$) | 27.00 [24.00, 28.70] | 23.20 [21.90, 25.55] | 25.65 [24.20, 27.90] | 27.90 [27.55, 28.30] | 24.55 [22.85, 26.60] |
| LVEF (%) | 67.5 [57.50, 70.00] | 64.0 [56.50, 69.00] | 59.5 [55.25, 62.50] | 28.0 [27.00, 28.50] | 64.0 [55.00, 70.00] |
| RWT | 0.50 [0.50, 0.61] | 0.36 [0.30, 0.41] | 0.35 [0.31, 0.38] | 0.30 [0.28, 0.31] | 0.79 [0.68, 0.91] |
| LVMI (gr.m$^2$) | 105.8 [90.90, 127.30] | 109.7 [97.55, 121.78 | 113.5 [93.55, 148.60] | 112.4 [101.2, 123.5] | 139.9 [132.12, 157.62] |
| AoS (cm$^2$) | 0.88 [0.80, 1.04] | 2.30 [2.00, 3.10] | 3.10 [2.70, 3.75] | _ | 3.00 [2.80, 3.40] |
| AoGrd (mmhg) | 44.00 [38.50, 53.25] | 4.40 [3.12, 6.75] | 8.80 [5.75, 10.45] | _ | 4.40 [4.30, 6.00] |
| Mitral EROA (cm$^2$) | _ | 0.44 [0.34, 0.66] | 0.08 [0.08, 0.08] | 0.12 [0.11, 0.13] | _ |
| Mtiral RV (ml) | _ | 68.50 [56.25, 102.75] | 15.0 [15.00, 15.00]" | 21.50 [18.75, 24.25]" | _ |
| Aortic EROA (cm$^2$) | _ | _ | 0.36 [0.32, 0.41] | _ | _ |
| Aortic RV (ml) | _ | _ | 67.0 [46.8, 87.5] | _ | _ |

Data are presented as median [first quartile - third quartile]; BMI: Body mass index, LVEF: left ventricular ejection fraction, RWT: relative wall thickness, LVMI: left ventricular mass index; AoS: aortic valve surface, AoGrd: aortic trans valvular mean systolic gradient, EROA: effective regurgitant orifice, RV: regurgitant volume.

**Table S3. Performance metrics of the surface motion camera for discriminating healthy participants from respiratory patients.**

| | **Area under the ROC curve** | **Youden index** | **Youden index coordinates** | **Sensitivity** | **Specificity** | **Accuracy** | **F1 score** |
|---|---|---|---|---|---|---|---|
| **Global analysis** | | | | | | | |
| Group 1 vs. Group 2 | 0.90 ± 0.07 | 0.708 | (1.0, 0.708) | 0.85 ± 0.10 | 0.74 ± 0.21 | 0.82 ± 0.07 | 0.87 ± 0.06 |
| **Subgroup analyses** | | | | | | | |
| Group 1 vs. Group 2-1 (mild) | 0.92 ± 0.09 | 0.773 | (0.833, 0.94) | 0.80 ± 0.32 | 0.84 ± 0.12 | 0.83 ± 0.12 | **0.66 ± 0.27** |
| Group 1 vs. Group 2-2 (moderate) | 0.89 ± 0.08 | 0.669 | (0.833, 0.836) | 0.87 ± 0.13 | 0.75 ± 0.21 | 0.81 ± 0.09 | 0.82 ± 0.08 |
| Group 1 vs. Group 2-3 (severe) | **0.96 ± 0.05** | 0.843 | (1.0, 0.843) | 0.86 ± 0.12 | 0.88 ± 0.12 | 0.87 ± 0.08 | 0.87 ± 0.08 |
| Kruskal-Wallis H<br>p-value | 13.7<br>0.0033 | – | – | 2.1<br>0.55 | 8.9<br>0.03 | 5.0<br>0.17 | 16.8<br>0.00076 |

Group 1: healthy participants (n=30);
Group 2: respiratory patient population (n=71);
Group 2-1: mild respiratory impairment (n=8);
Group 2-2: moderate respiratory impairment (n=32);
Group 2-3: severe respiratory impairment (n=31)(see Methods for subgroup definitions);
Youden index coordinates correspond to sensitivity and specificity at the optimal threshold derived from the ROC curve, whereas the values reported in the Sensitivity and Specificity columns correspond to fold-averaged estimates computed using a fixed decision threshold of 0.5.
F1 score: harmonic mean of precision and recall (sensitivity).
In columns with a significant Kruskal–Wallis effect, significant post hoc Dunn comparisons surviving multiple-comparison correction are indicated in bold.

.

**Table S4. Performance metrics of the surface motion camera for discriminating healthy participants from cardiac patients (acquisitions performed during free breathing).**

| | Area under the ROC curve | Youden index | Youden index coordinates | Sensitivity | Specificity | Accuracy | F1 score |
|---|---|---|---|---|---|---|---|
| **Global analyses** | | | | | | | |
| Group 1 (n=33) vs. Group 3 | 0.76 ± 0.10 | 0.435 | (1.0, 0.435) | **0.88 ± 0.08** | **0.39 ± 0.19** | 0.75 ± 0.08 | **0.84 ± 0.06** |
| **Subgroup analyses** | | | | | | | |
| Group 1 vs. Group 3-[1-2-3] | 0.78 ± 0.10 | 0.487 | (0.833, 0.654) | 0.81 ± 0.12 | 0.50 ± 0.18 | 0.71 ± 0.09 | 0.79 ± 0.07 |
| Group 1 vs. Group 3-[4-5] | 0.74 ± 0.13 | 0.390 | (1.0, 0.39) | **0.55 ± 0.26** | **0.81 ± 0.15** | 0.71 ± 0.10 | **0.56 ± 0.20** |
| Group 1 vs. Group 3.1 | **0.90 ± 0.10** | 0.709 | (1.0, 0.709) | 0.79 ± 0.18 | **0.83 ± 0.14** | 0.81 ± 0.12 | 0.80 ± 0.13 |
| Group 1 vs Group 3.3 | 0.75 ± 0.13 | 0.443 | (0.833, 0.609) | **0.64 ± 0.16** | **0.76 ± 0.15** | 0.70 ± 0.10 | **0.66 ± 0.12** |
| Kruskal-Wallis H<br>p-value | 28.3<br>$< 1\times10^{-4}$ | – | – | 42.6<br>$< 1\times10^{-7}$ | 65.7<br>$< 1\times10^{-12}$ | 17.6<br>0.001 | 50.6<br>$< 1\times10^{-9}$ |

Group 1: healthy participants (n=32);
Group 3: cardiac patient population (n=91);
Group 3.[1-2-3]: valvular heart disease (n=71);
Group 3.[4-5]: muscle heart disease (n=20);
Group 3.1: aortic stenosis (n=30);
Group 3.3: mitral regurgitation (n=28)
Youden index coordinates correspond to sensitivity and specificity at the optimal threshold derived from the ROC curve, whereas the values reported in the Sensitivity and Specificity columns correspond to fold-averaged estimates computed using a fixed decision threshold of 0.5.
F1 score: harmonic mean of precision and recall (sensitivity).
In columns with a significant Kruskal–Wallis effect, significant post hoc Dunn comparisons surviving multiple-comparison correction are indicated in bold.

**Table S5.** **Performance metrics of the surface motion camera for discriminating healthy participants from cardiac patients (acquisitions performed during breath-holding).**

| | **Area under the ROC curve** | **Youden index** | **Youden index coordinates** | **Sensitivity** | **Specificity** | **Accuracy** | **f1-score** |
|---|---|---|---|---|---|---|---|
| **Global analyses** | | | | | | | |
| Group 1 (n=32) vs. Group 3 | **0.78 ± 0.08** | 0.503 | (0.833, 0.67) | **0.92 ± 0.06** | **0.42 ± 0.16** | 0.79 ± 0.05 | **0.86 ± 0.03** |
| **Subgroup analyses** | | | | | | | |
| Group 1 vs. Group 3-[1-2-3] | 0.80 ± 0.12 | 0.519 | (0.833, 0.686) | 0.91 ± 0.08 | 0.50 ± 0.22 | 0.78 ± 0.09 | 0.85 ± 0.06 |
| Group 1 vs. Group 3-[4-5] | 0.72 ± 0.17 | 0.450 | (1.0, 0.45) | **0.58 ± 0.22** | **0.79 ± 0.17** | 0.71 ± 0.14 | **0.60 ± 0.19** |
| Group 1 vs. Group 3.1 | 0.89 ± 0.07 | 0.640 | (0.833, 0.807) | 0.83 ± 0.14 | **0.75 ± 0.18** | 0.79 ± 0.12 | 0.79 ± 0.11 |
| Group 1 vs Group 3.3 | 0.76 ± 0.12 | 0.481 | (1.0, 0.481) | **0.65 ± 0.20** | **0.74 ± 0.14** | 0.70 ± 0.10 | **0.65 ± 0.14** |
| Kruskal-Wallis H<br>p-value | 23.8<br>$< 1\times10^{-4}$ | – | – | 49.8<br>$< 1\times10^{-9}$ | 50.3<br>$< 1\times10^{-9}$ | 14.8<br>$5.1\times10^{-3}$ | 54<br>$< 1\times10^{-10}$ |

Group 1: healthy participants (n=32);
Group 3: cardiac patient population (n=91);
Group 3.[1-2-3]: valvular heart disease (n=71);
Group 3.[4-5]: muscle heart disease (n=20);
Group 3.1: aortic stenosis (n=30);
Group 3.3: mitral regurgitation (n=28)
Youden index coordinates correspond to sensitivity and specificity at the optimal threshold derived from the ROC curve, whereas the values reported in the Sensitivity and Specificity columns correspond to fold-averaged estimates computed using a fixed decision threshold of 0.5.
F1 score: harmonic mean of precision and recall (sensitivity).
In columns with a significant Kruskal–Wallis effect, significant post hoc Dunn comparisons are indicated in bold.

**Table S6.** **Summary of temporal synchronisation scores between gradient maxima and ECG signals across classification tasks and acquisition conditions.**

| Classification | Median | 70th percentile | 90th percentile |
|---|---|---|---|
| **cardiac surface motion camera (SMCc)** *acquisitions during free breathing* | | | |
| Group 1 vs. Group 3 | 0.76 | 0.89 | 0.99 |
| Group 1 vs Group 3.[1-2-3] | 0.82 | 0.92 | 0.98 |
| Group 1 vs Group 3.[4-5] | 0.78 | 0.89 | 0.98 |
| Group 1 vs Group 3.1 | 0.75 | 0.90 | 0.98 |
| Group 1 vs Group 3.3 | 0.81 | 0.90 | 0.98 |
| **cardiac surface motion camera (SMCc)** *acquisitions during breath-holding* | | | |
| Group 1 vs. Group 3 | 0.88 | 0.96 | 1.00 |
| Group 1 vs Group 3.[1-2-3] | 0.89 | 0.95 | 1.00 |
| Group 1 vs Group 3.[4-5] | 0.85 | 0.89 | 0.95 |
| Group 1 vs Group 3.1 | 0.85 | 0.96 | 1.00 |
| Group 1 vs Group 3.3 | 0.87 | 0.97 | 1.00 |

### Supplementary figures

**Figure S1. Schematic representation of the proposed network architectures:**
**A.** cardiac convolutional neural network; **B.** pulmonafry convolutional neural network.
N denotes the temporal dimension of the surface moction camera cardiac acquisitions (SMCc); CB, convolutional block; TFE, temporal feature extractor; C, classifier; and LSTM, long short-term memory.

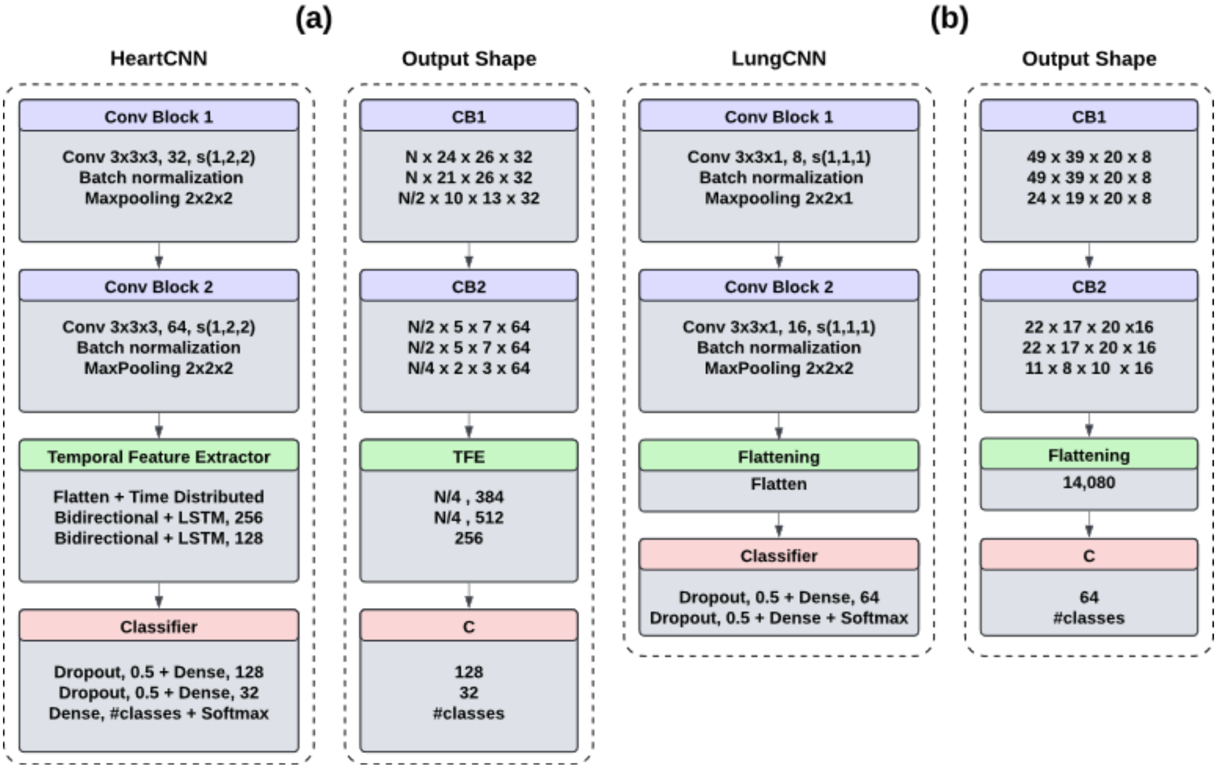

**Supplementary videos.**

*[as of April 27 2026, the supplementary videos can be downloaded at this temporary address : http://urlr.me/uEk6Nr]*

**Video S1.** Respiratory-related thoracic dynamic surface vibration map in a healthy participant (Group 1). The left panel ("*spatial slice*") shows the spatial admittance map at the frequency F. On the right, the top panel ("*frequential slice (F,X)*") shows the frequential slice of admittance map along Y axis, the middle panel ("*frequential slice (F,Y)*") shows the frequential slice of admittance map along X axis, the bottom panel ("*frequential signal along*" shows the frequential signal along (X,Y) pixel).
